# Beyond Six Feet: A Guideline to Limit Indoor Airborne Transmission of COVID-19

**DOI:** 10.1101/2020.08.26.20182824

**Authors:** Martin Z. Bazant, John W. M. Bush

## Abstract

The current revival of the world’s economy is being predicated on social distancing, specifically the Six-Foot Rule, a guideline that offers little protection from pathogen-bearing aerosol droplets sufficiently small to be continuously mixed through an indoor space. The importance of airborne transmission of COVID-19 is now widely recognized. While tools for risk assessment have recently been developed, no safety guideline has been proposed to protect against it. We here build upon models of airborne disease transmission in order to derive an indoor safety guideline that would impose an upper bound on the “cumulative exposure time”, the product of the number of occupants and their time in an enclosed space. We demonstrate how this bound depends on the rates of ventilation and air filtration, dimensions of the room, breathing rate, respiratory activity and face-mask use of its occupants, and infectiousness of the respiratory aerosols. By synthesizing available data from the best characterized indoor spreading events with respiratory drop-size distributions, we estimate an infectious dose on the order of ten aerosol-borne virions. The new virus is thus inferred to be an order of magnitude more infectious than its forerunner (SARS-CoV), consistent with the pandemic status achieved by COVID-19. Case studies are presented for classrooms and nursing homes, and a spreadsheet and online app are provided to facilitate use of our guideline. Implications for contact tracing and quarantining are considered, appropriate caveats enumerated. Particular consideration is given to respiratory jets, which may substantially elevate risk when face masks are not worn.

---

**C**oronavirus disease 2019 (COVID-19) is an infectious pneumonia that appeared in Wuhan, Hubei Province, China in December 2019 and has since caused a global pandemic (1, 2). The pathogen responsible for COVID-19, severe-acute-respiratory-syndrome coronavirus 2 (SARS-CoV-2), is known to be transported by respiratory droplets exhaled by an infected person (3–7). There are thought to be three primary routes of human-to-human transmission of COVID-19, large drop transmission from the mouth of an infected person to the mouth, nose or eyes of the recipient, physical contact with droplets deposited on surfaces (fomites) and subsequent transfer to the recipient’s respiratory mucosae, and inhalation of the microdroplets ejected by an infected person and held aloft by ambient air currents (6, 8). We subsequently refer to these three modes of transmission as, respectively, ‘large-drop’, ‘contact’ and ‘airborne’ transmission, while noting that the distinction between large-drop and airborne transmission is somewhat nebulous given the continuum of sizes of emitted droplets (9).^*^ We here build upon the existing theoretical framework for describing airborne disease transmission (12–18) in order to characterize the evolution of the concentration of pathogen-laden droplets in a well-mixed room, and the associated risk of infection to its occupants.

The Six-Foot Rule is a social-distancing recommendation by the U.S. Centers for Disease Control and Transmission, based on the assumption that the primary vector of pathogen transmission is the large drops ejected from the most vigorous exhalation events, coughing and sneezing (5, 19). Indeed, high-speed visualization of such events reveals that six feet corresponds roughly to the maximum range of the largest, millimeter-scale drops (20). Compliance to the Six-Foot Rule will thus substantially reduce the risk of such large-drop transmission. However, the liquid drops expelled by respiratory events are known to span a considerable range of scales, with radii varying from fractions of a micron to millimeters (9, 21).

There is now overwhelming evidence that indoor airborne transmission associated with relatively small, micron-scale aerosol droplets plays a dominant role in the spread of COVID-19 (4, 5, 7, 17–19, 22, 23), especially for so-called “super-spreading events” (24–27), which invariably occur indoors (28). For example, at the 2.5-hour-long Skagit Valley Chorale choir practice that took place in Washington State on March 10, some 53 of 61 attendees were infected, presumably not all of them within 6 feet of the initially infected individual (24). Similarly, when 23 of 68 passengers were infected on a 2-hour bus journey in Ningbo, China, their seated locations were uncorrelated with distance to the index case (27). Air-borne transmission was also implicated in the COVID-19 outbreak between residents of a Korean high-rise building whose apartments were linked via air ducts (29). Studies have also confirmed the presence of infectious SARS-CoV-2 virions in respiratory aerosols (30) suspended in air samples collected at distances as large as 16 feet from infected patients in a hospital room (3). Further evidence for the dominance of indoor airborne transmission has come from an analysis of 7324 early cases outside the Hubei Province, in 320 cities across mainland China (31). The authors found that all clusters of three or more cases occurred indoors, 80% arising inside apartment homes and 34% potentially involving public transportation; only a single transmission was recorded outdoors. Finally, the fact that face-mask directives have been more effective than either lock-downs or social distancing in controlling the spread of COVID-19 (22, 32) is consistent with indoor airborne transmission as the primary driver of the global pandemic.

The theoretical model developed herein informs the risk of airborne transmission resulting from the inhalation of small, aerosol droplets that remain suspended for extended periods within closed, well-mixed indoor spaces. When people cough, sneeze, sing, speak or breathe, they expel an array of liquid droplets formed by the shear-induced or capillary destabilization of the mucosal linings of the lungs and respiratory tract (8, 33, 34) and saliva in the mouth (35, 36). When the person is infectious, these droplets of sputum are potentially pathogen bearing, and represent the principle vector of disease transmission. The range of the exhaled pathogens is determined by the radii of the carrier droplets, which typically lie in the range of 0.1*µ*m - 1 mm. While the majority are submicron, the drop size distribution depends on the form of exhalation events (9). For normal breathing, the drop radii vary between 0.1 and 5.0 *µ*m, with a peak around 0.5 *µ*m (9, 37, 38). Relatively large drops are more prevalent in the case of more violent expiratory events such as coughing and sneezing (20, 39). The ultimate fate of the droplets is determined by their size and the air flows they encounter (40, 41). Exhalation events are accompanied by a time-dependent gas-phase flow emitted from the mouth that may be roughly characterized in terms of either continuous turbulent jets or discrete puffs (20, 37, 42). The precise form of the gas flow depends on the nature of the exhalation event, specifically the time-dependence of the flux of air expelled. Coughs and sneezes result in violent, episodic puff releases (20), while speaking and singing result in a puff train that may be well approximated as a continuous turbulent jet (37, 42). Eventually, the small droplets settle out of such turbulent gas flows. In the presence of a quiescent ambient, they then settle to the floor; however, in the well-mixed ambient more typical of a ventilated space, sufficiently small drops may be suspended by the ambient airflow and mixed throughout the room until being removed by the ventilation outflow or inhaled (see SI Appendix, Sec. 1A).

Existing theoretical models of airborne disease transmission in closed, well-mixed spaces are based on the seminal work of Wells (43) and Riley *et al*. (44), and have been applied to describe the spread of airborne pathogens including tuberculosis, measles, influenza, H1N1, coronavirus (SARS-CoV) (12–16, 45, 46) and most recently, the novel coronavirus (SARS-CoV-2) (17, 24). These models are all based on the premise that the space of interest is well mixed; thus, the pathogen is distributed uniformly throughout. In such well-mixed spaces, one is no safer from airborne pathogens at 60 feet than 6 feet. The Wells-Riley model (13, 15) highlights the role of the room’s ventilation outflow rate *Q* on the rate of infection, showing that the transmission rate is inversely proportional to *Q*, a trend supported by data on the spreading of airborne respiratory diseases on college campuses (47). The additional effects of viral deactivation, sedimentation dynamics and the polydispersity of the suspended droplets were considered by Nicas *et al*. (14) and Stilianakis & Drossinos (16). The equations describing pathogen transport in well-mixed, closed spaces are thus well established and have recently been applied to provide risk assessments for indoor airborne COVID-19 transmission (17, 18). We use a similar mathematical framework here in order to derive a simple safety guideline.

We begin by describing the dynamics of airborne pathogen in a well-mixed room, on the basis of which we deduce an estimate for the rate of inhalation of pathogen by its occupants. We proceed by deducing the associated infection rate from a single infected individual to a susceptible person. We illustrate how the model’s epidemiological parameter, a measure of the infectiousness of COVID-19, may be estimated from available epidemiological data, including transmission rates in a number of spreading events, and expiratory drop size distributions (9). Our estimates for this parameter are consistent with the pandemic status of COVID-19 in that they exceed those of SARS-CoV (17); however, our study calls for refined estimates through consideration of more such field data. Most importantly, our study yields a safety guideline for mitigating airborne transmission via limitation of indoor occupancy and exposure time, a guideline that allows for a simple quantitative assessment of risk in various settings. Finally, we consider the additional risk associated with respiratory jets, which may be considerable when face masks are not being worn.

## The Well-Mixed Room

We first characterize the evolution of the pathogen concentration in a well-mixed room. The assumption of well-mixedness is widely applied in the theoretical modeling of indoor airborne transmission (14, 16, 17), and its range of validity discussed in Section 1A of the SI Appendix. We describe the evolution of the airborne pathogen by adapting standard methods developed in chemical engineering to describe the “continuously stirred tank reactor” (48), as detailed in Section 1B of the SI Appendix. We assume that the droplet-borne pathogen remains airborne for some time before either being extracted by the room’s ventilation system, inhaled or sedimenting out. The fate of ejected droplets in a well-mixed ambient is determined by the relative magnitudes of two speeds, the settling speed of the drop in quiescent air, *v*_*s*_, and the ambient air circulation speed within the room, *v*_*a*_. Drops of radius *r* ≤100*µ*m and density *ρ*_*d*_ descend through quiescent air of density *ρ*_*a*_ and dynamic viscosity *μ*_*a*_ at the Stokes settling speed 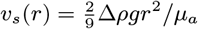, prescribed by the balance between gravity and viscous drag (49), where *g* is the gravitational acceleration and Δ *ρ* = *ρ*_*d*_ − *ρ*_*a*_.

We consider a well-mixed room of area *A*, depth *H* and volume *V* = *HA* with ventilation outflow rate *Q* and outdoor air change rate (typically reported as air changes per hour, or ACH) *λ*_*a*_ = *Q/V*. In the case of mechanical ventilation, there is an additional recirculation flow rate *Q*_*r*_ that further contributes to the well-mixed state of the room, but alters the emergent drop-size distributions only if accompanied by filtration. The mean air velocity, *v*_*a*_ = (*Q* + *Q*_*r*_)*/A*, prescribes the air mixing time, *τ*_*a*_ = *H/v*_*a*_ = *H*^2^*/*(2*D*_*a*_), where *D*_*a*_ = *v*_*a*_*H/*2 is the turbulent diffusivity defined in terms of the largest eddies (50, 51), those on the scale of the room (52). The timescale of the droplet settling from a well-mixed ambient corresponds to that through a quiescent ambient (50, 51, 53), as justified in SI Appendix, Sec.1B. Equating the characteristic times of droplet settling, *H/ v*_*s*_, and removal, *V /Q*, indicates a critical drop radius 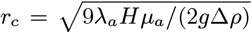 above which drops generally sediment out, and below which they remain largely suspended within the room prior to removal by ventilation outflow. We here define airborne transmission as that associated with droplets with radius *r < r*_*c*_. The relevant physical picture, of particles settling from a well-mixed environment, is commonly invoked in the contexts of stirred aerosols (50) and sedimentation in geophysics (53). The additional effects of ventilation, particle dispersity and pathogen deactivation in the context of airborne disease transmission were considered by Nicas *et al*. (14), Stiliankakis & Drossinos (16) and Buonanno et al. (17, 18), whose models will be built upon here.

In Sec.1A of SI Appendix, we provide justification for our assumption of the well-mixed room. It is noteworthy that, even in the absence of forced ventilation, there will generally be some mixing in an enclosed space: natural ventilation will lead to flows through windows and doors, as well as leakage though construction materials and joints. Moreover, occupants serve to enhance air flow through their motion and respiration. Traditionally, ventilation standards for American homes (ASHRAE) recommend a minimal outdoor air exchange rate of *λ*_*a*_ = 0.35/h, a value comparable to the average of 0.34/h reported for Chinese apartments, including winter in Wuhan during the initial outbreak (54). Even with such minimal ventilation rates, for a room of height *H* = 2.1m there is an associated critical drop size of radius *r*_*c*_ = 1.3*µ*m. In order to guard against infectious aerosols, ASHRAE now recommend ventilation rates greater than *λ*_*a*_ = 6*/h*, which corresponds to *r*_*c*_ = 5.5*µ*m. The “airborne” droplets of interest here, those of radius *r < r*_*c*_, thus constitute a significant fraction of those emitted in most respiratory events (9, 23, 37).

Wells (55) argued that exhaled drops with diameter less than around 100 *µ*m will nearly evaporate before settling. The resulting “droplet nuclei” consist of residual solutes, including dissolved salts, carbohydrates, proteins and pathogens, which are typically hygroscopic and retain significant quantities of bound water (56, 57). For a droplet with initial radius *r*_0_, the equilibrium size, 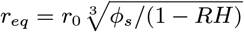, is reached over an evaporation time scale, 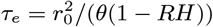, where *ϕ*_*s*_ is the initial solute volume fraction, *RH* is the relative humidity, and *θ* = 4.2 10^−10^ m^2^/s at 25 ^*°*^C (57). In dry air (*RH* ≪1), saliva droplets, which typically contain 0.5% solutes and a similar volume of bound water (*ϕ*_*s*_ ≈ 1%), can thus lose up to 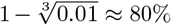 of their initial size (57). Conversely, droplets of airway mucus shrink by as little as 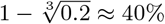, since they typically contain 5-10% gel-forming mucins (glycosylated proteins) and comparable amounts of bound water (58). The evaporation time at 50% RH ranges from *τ*_*e*_ = 1.2 ms for *r*_0_ = 0.5*µ*m to 12s at 50*µ*m. These inferences are consistent with experiments demonstrating that stable respiratory aerosol distributions in the range *r*_*eq*_ *<* 10*µ*m are reached within 0.8s of exhalation (9). While we note that the drop size distributions will in general depend on the relative humidity, we proceed by employing the equilibrium drop distributions measured directly (9, 37).

We consider a polydisperse suspension of exhaled droplets characterized by the number density *n*_*d*_(*r*) (per volume of air, per radius) of drops of radius *r* and volume 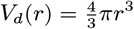. The drop size distribution *n*_*d*_(*r*) is known to vary strongly with respiratory activity and various physiological factors (9, 17, 38). The drops contain a microscopic pathogen concentration *c* _*v*_ (*r*), a drop-size-dependent probability of finding individual virions (3, 30, 59), usually taken to be that in the sputum (RNA copies per mL) (17, 60).

The virions become deactivated (non-infectious) at a rate *λ*_*v*_ (*r*) that will in general depend on droplet radius, temperature and humidity (61). Using data for human influenza viruses (62), a roughly linear relationship between *λ*_*v*_ and *RH* can be inferred (61, 63), which provides rationale for the seasonal variation of flu outbreaks, specifically the decrease from humid summers to dry winters. Recent experiments on the aerosol viability of model viruses (bacteriophages) by Lin and Marr (64) have further revealed a non-monotonic dependence of *λ*_*v*_ on relative humidity. Specifically, the deactivation rate peaks at intermediate values of relative humidity, where the cumulative exposure of virions to disinfecting salts and solutes is maximized. Since the dependence *λ*_*v*_ (*RH*) is not yet well characterized experimentally for SARS-CoV-2, we follow Miller *et al*. (24) and treat the deactivation rate as bounded by existing data, specifically, *λ*_*v*_ = 0 (no deactivation measured in 16 hours at 22 ± 1 ^*°*^*C* and *RH* = 53 *±* 11% (65)) and *λ*_*v*_ = 0.63/h (corresponding to a half life of 1.1h at 23 ± 2 ^*°*^*C* and *RH* = 65% (66)). Pending further data for SARS-CoV-2, we assume *λ*_*v*_ = 0.6*RH*, and note the rough consistency of this estimate with that for MERS-CoV (Middle East Respiratory Syndrome coronavirus) at 25 ^*°*^*C* and *RH* = 79% (67), specifically *λ*_*v*_ = 1.0*/*h. Finally, we note that the effective viral deactivation rates may be enhanced by use of either ultraviolet radiation (UV-C) (68) or chemical disinfectants (*e*.*g*. H_2_O_2_, O_3_) (69).

The influence of air filtration and droplet settling in ventilation ducts may be incorporated by augmenting *λ*_*v*_ (*r*) by an amount *λ*_*f*_ (*r*)= *p*_*f*_ (*r*) *λ*_*r*_, where *p*_*f*_ (*r*) is the probability of droplet filtration and *λ*_*r*_ = *Q*_*r*_ */V*. The recirculation flow rate, *Q*_*r*_, is commonly expressed in terms of the primary outdoor air fraction, *Z*_*p*_ = *Q/*(*Q*_*r*_ + *Q*), where *Q* + *Q*_*r*_ is the total air flow rate. We note that the United States Environmental Protection Agency defines high-efficiency particulate air (HEPA) filtration (70) as that characterized by *p*_*f*_ *>* 99.97% for aerosol particles. Ordinary air filters have required Minimum Efficiency Reporting Value (MERV) ratings of *p*_*f*_ = 20 − 90% in specific size ranges. Other types of filtration devices (22), such as electrostatic precipitators (71) with characteristic *p*_*f*_ values of 45 − 70%, can also be included in this framework.

We seek to characterize the concentration *C*(*r, t*) (specifically, number/volume per radius) of pathogen transported by drops of radius *r*. We assume that each of *I*(*t*) infectious individuals exhales pathogen-laden droplets of radius *r* at a constant rate *P* (*r*)= *Q*_*b*_*n*_*d*_(*r*)*V*_*d*_(*r*)*p*_*m*_(*r*)*c*_*v*_(*r*) (number/time per radius), where *Q*_*b*_ is the breathing flow rate (exhaled volume per time). We introduce a mask penetration factor, 0 *< p*_*m*_(*r*) *<* 1, that roughly accounts for the ability of masks to filter droplets as a function of drop size (72–75).^†^ The concentration, *C*(*r, t*), of pathogen suspended within drops of radius *r* then evolves according to

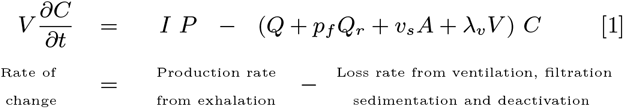

where *v*_*s*_(*r*) is the particle settling speed and *p*_*f*_ (*r*) is again the probability of drop filtration in the recirculation flow *Q*_*r*_. Owing to the dependence of the settling speed on particle radius, the population of each drop size evolves, according to equation [1], at different rates. Two limiting cases of Eq. [1] are of interest. For the case of *λ* _*v*_ = *v*_*s*_ = *Q*_*r*_ = 0, drops of infinitesimal size that are neither deactivated nor removed by filtration, it reduces to the Wells-Riley model (43, 44). For the case of *λ*_*v*_ = *P* = *Q* = *Q*_*r*_ = 0, a non-reacting suspension with no ventilation, it corresponds to established models of sedimentation from a well-mixed ambient (50, 53). For the sake of notational simplicity, we define a size-dependent sedimentation rate *λ*_*s*_(*r*)= *v*_*s*_(*r*)*/H* = *λ*_*a*_(*r/r*_*c*_)^2^ as the inverse of the time taken for a drop of radius *r* to sediment from ceiling to floor in a quiescent room.

When one infected individual enters a room at time *t* = 0, so that *I*(0) = 1, the radius-resolved pathogen concentration increases as, 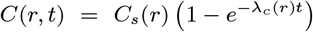, relaxing to a steady value, *C*_*s*_(*r*) = *P* (*r*)*/*(*λ*_*c*_(*r*)*V*), at a rate, *λ*_*c*_(*r*)=*λ*_*a*_ +*λ*_*f*_ (*r*)+*λ*_*s*_(*r*)+*λ*_*v*_(*r*). Note that both the equilibrium concentration and the timescale to approach it are decreased by the combined effects of ventilation, air filtration, particle settling and deactivation (14, 63). Owing to the dependence of this adjustment process on the drop size, one may understand it as a dynamic sifting process wherein larger droplets settle out and reach their equilibrium concentration relatively quickly. However, we note that, in the absence of filtration and deactivation (*λ*_*f*_ = *λ*_*v*_ = 0), the adjustment time, 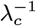, depends only weakly on drop size, varying from *V/*(2*Q*) for the largest airborne drops (with radius *r*_*c*_) to *V /Q* for infinitesimal drops. The sedimentation rate of the ‘airborne’ droplets of radius *r ≤ r*_*c*_ is thus bounded above by the air exchange rate, *λ*_*s*_(*r*) ≤ *λ*_*a*_. The exhaled drop-size distribution depends strongly on respiratory activity (9, 17, 37, 38); thus, so too must the radius-resolved concentration of airborne pathogen. The predicted dependence on respiratory activity (9) of the steady-state volume fraction of airborne droplets, *ϕ* (*r*) = *C*_*s*_(*r*)*/c*_*v*_ (*r*), is illustrated in Fig. 1.

**Fig. 1.**
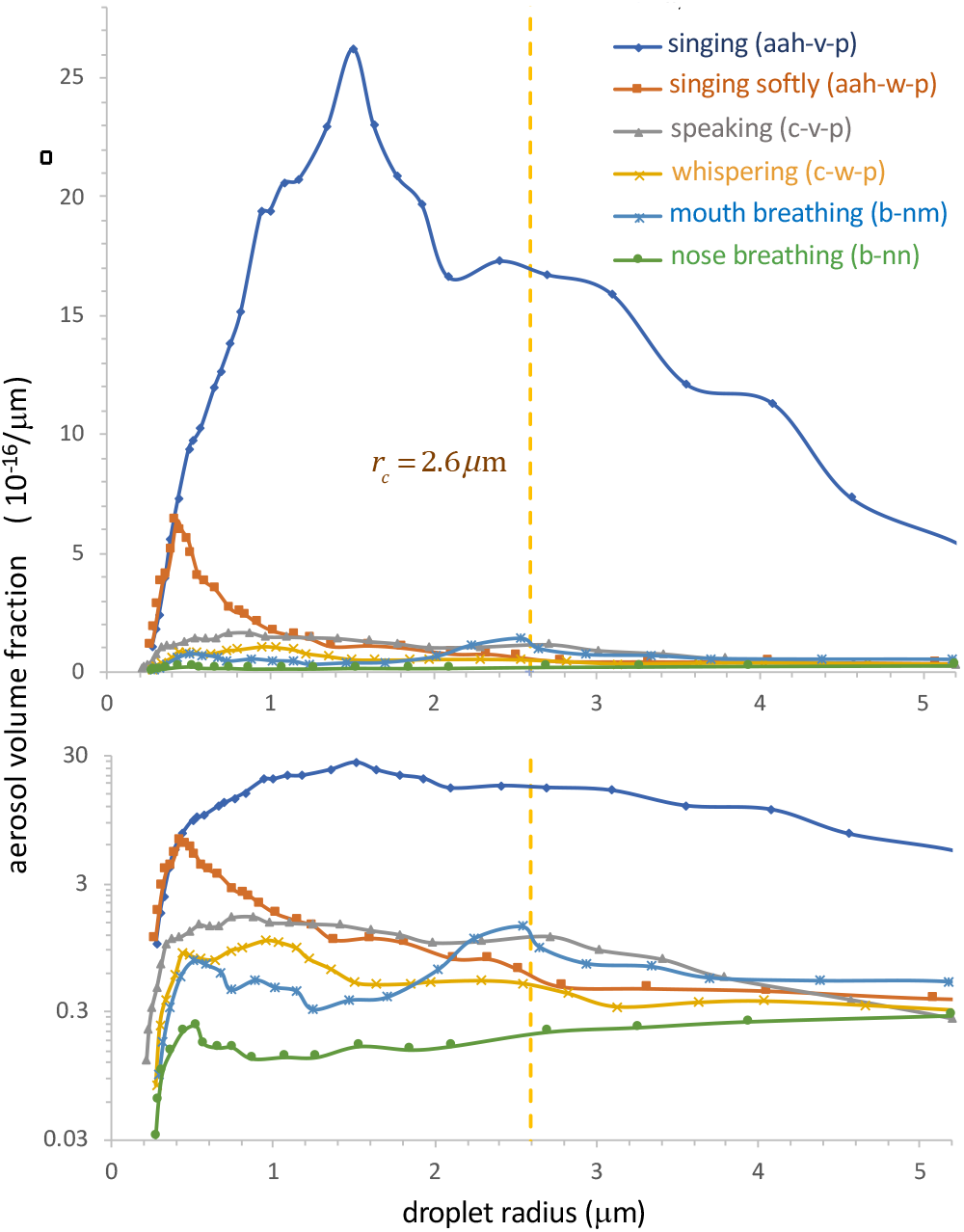
Model predictions for the steady-state, droplet-radius-resolved aerosol volume fraction, *ϕ*_*s*_(*r*), produced by a single infectious person in a well-mixed room. The model accounts for the effects of ventilation, pathogen deactivation and droplet settling for several different types of respiration in the absence of face masks (*p*_*m*_ = 1). The ambient conditions are taken to be those of the Skagit Valley Chorale super-spreading incident (24, 26) (*H* = 4.5m, *A* = 180m^2^ λ _*a*_ = 0.65h^−1^, *r*_*c*_ = 2.6*µ*m, *λ*_*v*_ = 0.3h^−1^, *RH* = 50%). The expiratory droplet size distributions are computed from the data of Morawska *et al*. at *RH* = 59.4%(9) (see their Fig. 3) for aerosol concentration per log-diameter, using *n*_*d*_(*r*) = (*dC/d* log *D*)*/*(*r* ln 10). The breathing flow rate is assumed to be 0.5 m^3^ /h for nose and mouth breathing, 0.75 m^3^ /h for whispering and speaking, and 1.0 m^3^ /h for singing.

We define the airborne disease transmission rate, *β*_*a*_(*t*), as the mean number of transmissions per time per infectious individual per susceptible individual. One expects *β*_*a*_(*t*) to be proportional to the quantity of pathogen exhaled by the infected person, and to that inhaled by the susceptible person. Gammaitoni and Nucci (12) defined the airborne transmission rate as *β*_*a*_(*t*) = *Q*_*b*_*c*_*i*_*C*_*s*_(*t*) for the case of a population evolving according to the Wells-Riley model and inhaling a monodisperse suspension. Here, *c*_*i*_ is the viral infectivity, the parameter that connects the fluid physics to the epidemiology, specifically the concentration of suspended pathogen to the infection rate. We note its relation to the notion of “infection quanta” in the epidemiological literature (43). Specifically, *c*_*i*_ *<* 1 is the infection quanta per pathogen, while 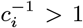 is the “infectious dose”, the number of aerosol-borne virions required to cause infection with probability 1 − *e*^−1^ = 63%.

For the polydisperse suspension of interest here, we define the airborne transmission rate as

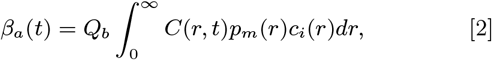

thereby accounting for the protective properties of masks, and allowing for the possibility that the infectivity *c*_*i*_(*r*) depends on droplet size. Different droplet sizes may emerge from, and penetrate into, different regions of the respiratory tract (33, 36, 78), and so have different *c*_*i*_(*r*); moreover, virions in relatively small droplets may diffuse to surfaces more rapidly and so exchange with bodily fluids more effectively. Such a size dependence in infectivity, *c*_*i*_(*r*), is also consistent with reports of enhanced viral shedding in micron-scale aerosols compared to larger drops for both influenza virus (59) and SARS-CoV-2 (30).

## Indoor Safety Guideline

The reproduction number of an epidemic, ℛ_0_, is defined as the mean number of transmissions per infected individual. Provided ℛ_0_ *<* 1, a disease will not spread at the *population* level (79). Estimates of ℛ_0_ for COVID-19 have been used to compare its rate of spread in different regions and its dependence on different control strategies (32, 80–82) and, most recently, viral variants (83, 84). We here define an analogous reproductive number for indoor, airborne transmission, ℛ_*in*_(*τ*), as the expected number of transmissions in a room of total occupancy *N* over a time *τ* from a single infected person entering at *t* = 0.

Our safety guideline sets a small risk tolerance *ϵ* (typically 1-10%) for the indoor reproductive number, defined as

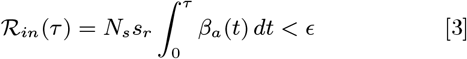

The number of susceptibles, *N*_*s*_ = *p*_*s*_(*N* − 1), may include all others in the room (*p*_*s*_ = 1), or be reduced by the susceptible probability *p*_*s*_ *<* 1, the fraction of the local population not yet exposed or immunized. The relative transmissibility (or susceptibility), *s*_*r*_ allows for consideration of different transmission rates for different sub-populations or viral strains.

In the limit of *ϵ* ≪ 1, one may interpret ℛ_*in*_(*τ*) as the probability of the first transmission, which is approximately equal to the sum of the *N*_*s*_ independent probabilities of transmission to any particular susceptible individual in a well-mixed room^‡^. In SI Appendix Sec. 3, we show that this guideline follows from standard epidemiological models, including the Wells-Riley model, but note that it has broader generality. The exact transient safety bound appropriate for the time-dependent situation arising directly after an infected index case enters a room, is evaluated in SI Appendix, Sec. 2.

We here focus on a simpler and more conservative guideline that follows for long times relative to the air residence time, 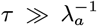 (which may vary from minutes to hours, and is necessarily greater than *λ*_*c*_(*r*)^−1^), when the airborne pathogen has attained its equilibrium concentration *C*(*r, t*) → *C*_*s*_(*r*). In this equilibrium case, the transmission rate (2) becomes

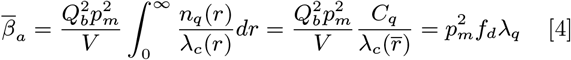

where, for the sake of simplicity, we assume constant mask filtration *p*_*m*_ over the entire range of aerosol drop sizes. We define the microscopic concentration of infection quanta per liquid volume as *n*_*q*_(*r*) = *n*_*d*_(*r*)*V*_*d*_(*r*)*c*_*v*_(*r*)*c*_*i*_(*r*), and the concentration of infection quanta or “infectiousness” of exhaled air, 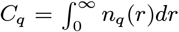. The latter is the key disease-specific parameter in our model, which can also be expressed as the rate of quanta emission, *λ*_*q*_ = *Q*_*b*_*C*_*q*_, by an infected person. The second equality in Eq. [4] defines the effective infectious drop radius 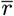, given in Eq. [S7]. The third equality defines the dilution factor, 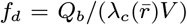, the ratio of the concentration of infection quanta in the well-mixed room to that in the unfiltered breath of an infected person. As we shall see in what follows, this dilution factor provides a valuable diagnostic in assessing the relative risk of various forms of exposure.

We thus arrive at a simple guideline, appropriate for steady-state situations, that bounds the *cumulative exposure time* (CET):

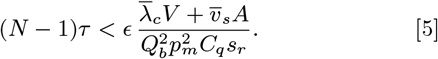

where 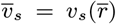 and 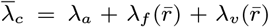 is the effective air exchange rate accounting for air filtration and virus deactivation. The effect of relative humidity on the droplet size distribution can be captured by multiplying 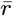 by 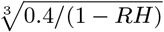, since the droplet distributions used in our analysis were measured at *RH* = 60% (9).

By noting that the sedimentation rate of aerosols is usually less than the air exchange rate, *λ*_*s*_(*r*) *< λ*_*a*_, and by neglecting the influence of both air filtration and pathogen deactivation, we deduce from Eq. [5] a more conservative bound on the CET,

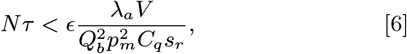

the interpretation of which is immediately clear. To minimize risk of infection, one should avoid spending extended periods in highly populated areas. One is safer in rooms with large volume and high ventilation rates. One is at greater risk in rooms where people are exerting themselves in such a way as to increase their respiration rate and pathogen output, for example by exercising, singing or shouting. Since the rate of inhalation of contagion depends on the volume flux of both the exhalation of the infected individual and the inhalation of the susceptible person, the risk of infection increases as 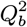. Likewise, masks worn by both infected and susceptible persons will reduce the risk of transmission by a factor 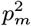, a dramatic effect given that *p*_*m*_ ≤ 0.1 for moderately high quality masks (73, 74).

## Application to COVID-19

The only poorly constrained quantity in our guideline is the epidemiological parameter, *C*_*q*_*s*_*r*_, the product of the concentration of exhaled infection quanta by an infectious individual, *C*_*q*_, and the relative transmissibility, *s*_*r*_. We emphasize that *C*_*q*_ and *s*_*r*_ are expected to vary widely between different populations (85–90), among individuals during progression of the disease (91, 92) and between different viral strains (83, 84). Nevertheless, we proceed by making rough estimates for *C*_*q*_ for different respiratory activities on the basis of existing epidemiological data gathered from early super-spreading events of COVID-19. Our inferences provide a baseline value for *C*_*q*_, relevant for elderly individuals exposed to the original strain of SARS-CoV-2, that we may rescale by the relative transmissibilty *s*_*r*_ in order to consider different populations and viral strains. We make these inferences with the hope that such an attempt will motivate the acquisition of more such data, and so to improved estimates for *C*_*q*_ and *s*_*r*_ for different populations in various settings.

An inference of *C*_*q*_ = 970 quanta/m^3^ was made by Miller *et al*. (24) in their recent analysis of the Skagit Valley Chorale super-spreading incident (26), on the basis of the assumption that the transmission was described in terms of the Wells-Riley model (12, 13, 17, 44). To be precise, they inferred a quanta emission rate of 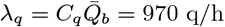 for a mean breathing rate of 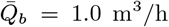 apprpriate for singing (24). This inference is roughly consistent with studies of other related viral diseases. For example, Liao *et al*. (45) estimated *C*_*q*_ = 28 quanta/m^3^ from the rate of indoor spreading of SARS-CoV, in a hospital and an elementary school. Estimates of *C*_*q*_ for H1N1 influenza fall in the range 15 − 128 quanta/m^3^ (46). For SARS-CoV-2, Buonanno *et al*. (17) estimate a *C*_*q*_ range of 10.5-1030 quanta/m^3^, on the basis of the estimated infectivity *c*_*i*_ = 0.01 − 0.1 of SARS-CoV (93) and the reported viral loads in sputum (91, 92, 94), and note that the precise value depends strongly on the infected person’s respiratory activity. Notably, their range spans the high value inferred for the Skagit Valley Chorale (24), and all of our inferences to follow.

We proceed by estimating quanta concentrations, *C*_*q*_, or equivalently, quanta emission rates, *λ*_*q*_ = *Q*_*b*_*C*_*q*_, for different forms of respiration. First, we solve Eq. [1] to obtain the steady-state radius-resolved droplet volume fraction *ϕ*_*s*_(*r*) for various hypothetical expiratory activities in the room of the Skagit Valley Chorale, using the drop size distributions of Morawska *et al*. (9). Our results are shown in Fig. 1. Integrating each curve up to the critical radius *r*_*c*_, we then obtain an activity-dependent volume fraction of infectious airborne droplets 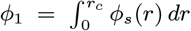 in the choir room (see SI Appendix). Finally, we assume that the inferred value, *C*_*q*_ = 970 quanta/m^3^, for the super-spreading incident (24) that resulted from the expiratory activity most resembling singing (voiced “aahs” with pauses for recovery (9)), and deduce values of *C*_*q*_ for other forms of respiration by rescaling with the appropriate *ϕ*_1_ values. Our predictions for the dependence of *C*_*q*_ on respiratory activity are shown in Fig. 2. For validation, we also show estimates for *C*_*q*_ based on the recent measurements of activity-dependent aerosol concentrations reported by Asadi *et al*. (37, 38). Specifically, we calculated the aerosol volume fractions from the reported drop-size distributions (from Fig. 5 of Ref.(38)) for a different set of expiratory activities that included various breathing patterns and speaking aloud at different volumes. We then used these volume fractions to rescale the value *C*_*q*_ = 72 quanta/m^3^ for speaking at intermediate volume (38), which we chose to match the value inferred for the most similar respiratory activity considered by Morawska *et al*. (9), specifically voiced counting with pauses (9). Notably, the quanta concentrations so inferred, *C*_*q*_, are consistent across the full range of activities, from nasal breathing at rest (1-10 q/m^3^) to oral breathing and whispering (5-40 q/m^3^), to loud speaking and singing (100-1000 q/m^3^).

**Fig. 2.**
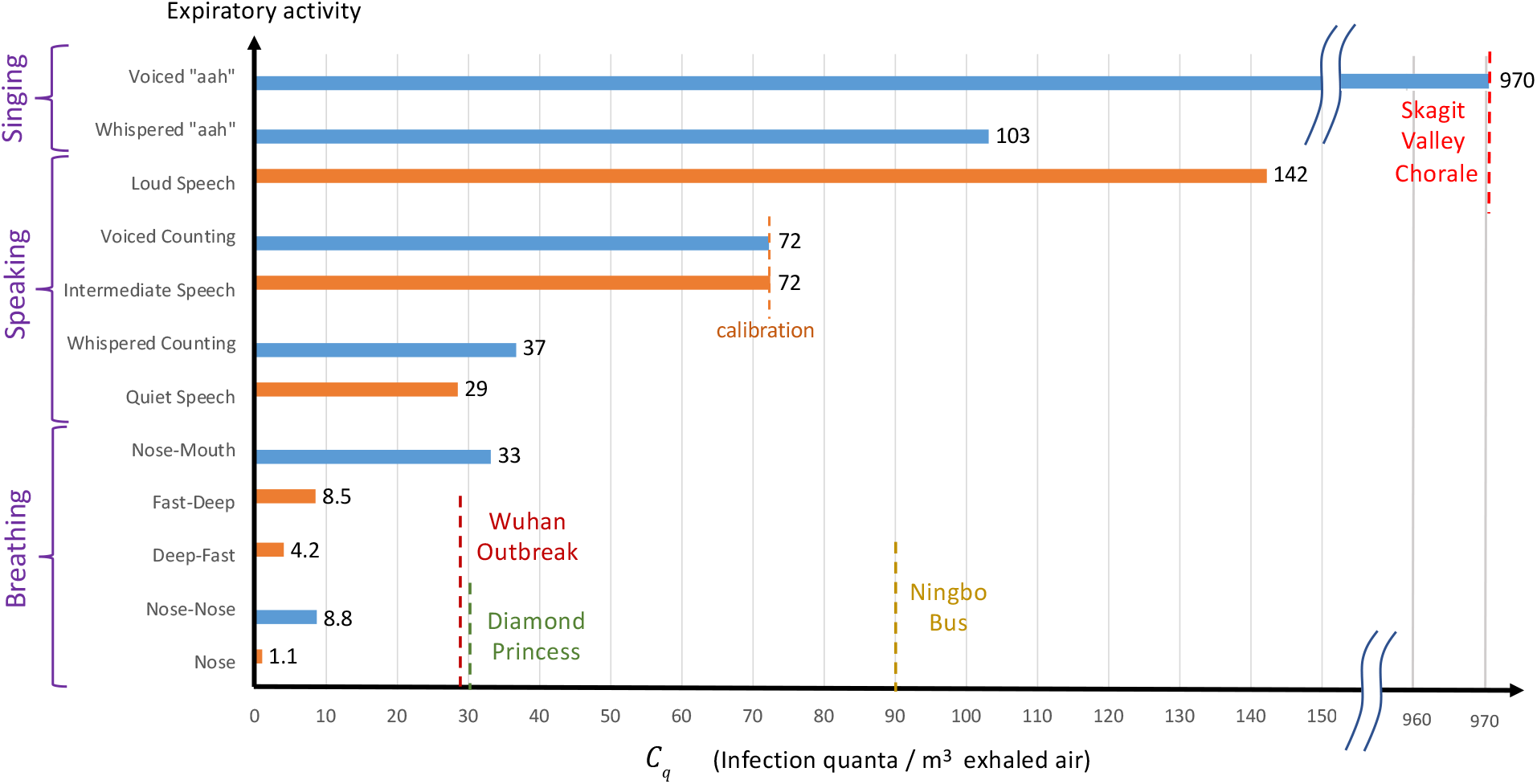
Estimates of the “infectiousness” of exhaled air, *C*_*q*_, defined as the peak concentration of COVID-19 infection quanta in the breath of an infected person, for various respiratory activities. Values are deduced from the drop-size distributions reported by Morawska *et al*. (9) (blue bars) and Asadi *et al*. (38) (orange bars). The only value reported in the epidemiological literature, *C*_*q*_ = 970 quanta/m^3^, was estimated (24) for the Skagit Valley Chorale super-spreading event (26), which we take as a baseline case (*s*_*r*_ = 1) of elderly individuals exposed to the original strain of SARS-CoV-2. This value is rescaled by the predicted infectious aerosol volume fractions,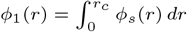 obtained by integrating the steady-state size distributions reported in Fig. 1 for different expiratory activities (9). Aerosol volume fractions calculated for various respiratory activities from Fig. 5 of Asadi et al. (38) are rescaled so that the value *C*_*q*_ = 72 quanta/m^3^ for “intermediate speaking” matches that inferred from Morawska *et al*.’s (9) for “voiced counting”. Estimates of *C*_*q*_ for the outbreaks during the quarantine period of the Diamond Princess (25) and the Ningbo bus journey (27), as well as the initial outbreak in Wuhan City (2, 80) are also shown (see SI Appendix for details).

Our inferences for *C*_*q*_ from a number of super-spreading events are also roughly consistent with physiological measurements of viral RNA in the bodily fluids of COVID-19 patients at peak viral load. Specifically, our estimate of *C*_*q*_ = 72 quanta/m^3^ for voiced counting (9) and intermediate-volume speech (38) with integrated aerosol volume fractions *ϕ*_1_ = 0.36 and 0.11 (*µ*m/cm)^3^ corresponds, respectively, to microscopic concentrations of *c*_*q*_ = *c*_*i*_*c*_*v*_ =2 × 10^8^ and 7 × 10^8^ quanta/mL (see SI Appendix). Respiratory aerosols mainly consist of sputum produced by the fragmentation (95) of mucous plugs and films in the bronchioles and larynx (33–35). Larger droplets are thought to form by fragmentation of saliva in the mouth (35, 36). Airborne viral loads are usually estimated from that of saliva or sputum (60, 91, 92, 94, 96). After incubation, viral loads, *c* _*v*_, in sputum tend to peak in the range 10^8^ − 10^11^ RNA copies/mL (91, 92, 94), while much lower values have been reported for other bodily fluids (91, 92, 97). Virus shedding in the pharynx remains high during the first week of symptoms and reaches 7 × 10^8^ RNA copies per throat swab (91) (typically 1-3 mL). Since viral loads are 20-50% greater in sputum than in throat swabs (92), the most infectious aerosols are likely to contain *c*_*v*_ ≈ 10^9^ RNA copies/mL. Using this viral load and assuming *c*_*i*_ = 2% based on previous inferences for SARS-CoV (93), Buonanno *et al*. (17) estimated *c*_*q*_ = 2 × 10^7^ q/mL for SARS-CoV-2, an order of magnitude below our inferences obtained directly from spreading data for COVID-19 (9, 38). The inference that SARS-CoV-2 is ten times more infectious than SARS-CoV, with *c*_*i*_ ∼ 10% (an infectious dose on the order of ten aerosol-borne virions), is consistent with the fact that only the former caused a pandemic.

Our findings are consistent with emerging virological (3, 30, 65, 66) and epidemiological (5, 19, 23, 27, 28) evidence that SARS-CoV-2 is present and extremely infectious in respiratory aerosols and that indoor airborne transmission is the dominant driver of the COVID-19 pandemic (4, 22). Further support for this hypothesis is provided by crudely applying our indoor transmission model to a number of slightly less well characterized spreading events, as detailed in the SI Appendix, all of which yield roughly consistent values of *C*_*q*_ (shown in Fig. 2). For the initial outbreak of COVID-19 in Wuhan City (2, 80), we assume that spreading occurred predominantly in family apartments, as is consistent with the inference that 80% of transmission clusters arose in people’s homes (31). We may then tentatively equate the average reproduction number estimated for the Wuhan outbreak (80), ℛ_0_ = 3.3, with the indoor reproduction number, ℛ_*in*_(*τ*). We use *τ*= 5.5 days as the exposure time, assuming that it corresponds to mean time before the onset of symptoms and patient isolation. We consider the mean household size of 3.03 persons in a typical apartment with area 30 m^2^/person and a winter bedroom ventilation rate of 0.34 ACH (54), and assume that *λ*_*v*_ = 0.3/h and 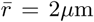. We thus infer *C*_*q*_ = 30 quanta/m^3^, a value expected for normal breathing (see Fig. 2).

For the Ningbo bus incident all model parameters are known except for the air exchange rate. We estimate *λ*_*a*_ = 1.25/h for a moving bus with closed windows, based on studies of pollutants in British transit buses (98). We thus infer *C*_*q*_ = 90 q/m^3^, a value that lies in the range of intermediate speaking, as might be expected on board a bus filled to capacity. Considering the uncertainty in *λ*_*a*_, one might also infer a value consistent with resting on a quiet bus; in particular, choosing *λ*_*a*_ = 0.34/h yields *C*_*q*_ = 57 q/m^3^. Finally, we infer a value of *C*_*q*_ = 30 q/m^3^ from the spreading event on board the quarantined Diamond Princess cruise ship (25), a value consistent with the passengers being primarily at rest. However, we note that the extent to which the Diamond Princess can be adequately described in terms of a well-mixed space remains the subject of some debate (see SI Appendix, Sec. 5A).

We proceed by making the simplifying assumption that the dependence of *C*_*q*_ on expiratory activity illustrated in Fig. 2 is universal, but retain the freedom to rescale these values by the relative transmissibility *s*_*r*_ for different age groups and viral strains. It is well established that children have considerably lower hospitalization and death rates (85–87), but there is growing evidence that they also have lower transmissibility (88– 90, 99, 100). A recent study of household clusters suggests that children are rarely index cases or involved in secondary transmissions (88). The best controlled comparison comes from quarantined households in China, where social contacts were reduced 7-8 fold during lockdowns (100). Compared to the elderly (over 65 years old) for which we have assigned *s*_*r*_ = 1, the relative susceptibility of adults (aged 15-64) was found to be *s*_*r*_ = 68%, while that of children (aged 0-14) was *s*_*r*_ = 23%. We proceed by using these values of *s*_*r*_ for these three different age groups and the original strain of SARS-CoV-2 in our case studies. However, we anticipate the need to revise these *s*_*r*_ values for new viral variants, such as the lineage B.1.1.7 (VOC 202012/01) (83, 84), which recently emerged in the United Kingdom with 60% greater transmissibility and elevated risk of infection among children.

In summary, our inferences of *C*_*q*_ and *s*_*r*_ from a diverse set of indoor spreading events and from independent physiological data are sufficiently self-consistent to indicate that the values reported in Fig. 2 may prove to be sufficient to apply the safety guideline in a quantitative fashion. Our hope is that our attempts to infer *C*_*q*_ will motivate the collection of more such data from spreading events, which might then be used to refine our necessarily crude initial estimates.

## Case studies

We proceed by illustrating the value of our guideline in estimating the maximum occupancy or exposure time in two settings of particular interest, the classroom and an elder care facility. Considering our inferences from the data and the existing literature, it would appear reasonable to illustrate our guideline for COVID-19 with the conservative choice of *C*_*q*_ = 30 quanta/m^3^. However, we emphasize that this value is expected to vary strongly with different demographics and respiratory activity levels (17). In taking the value of *C*_*q*_ = 30 quanta/m^3^, we are assuming that in both settings considered, occupants are engaged in relatively mild respiratory activities consistent with quiet speech or rest. In assessing critical cumulative exposure times for given populations, we stress that the tolerance *ϵ* is a parameter that should be chosen judiciously according to the vulnerability of the population, which varies dramatically with age and preexisting conditions (85–88).

We first apply our guideline to a typical classroom in the United States, designed for the standard occupancy of 19 students and their teacher (see Fig. 3(a)). The importance of adequate ventilation and mask use is made clear by our guideline. For normal occupancy and without masks, the safe time after an infected individual enters the classroom is 1.2 hours for natural ventilation and 7.2 hours with mechanical ventilation, according to the transient bound, Eq. [S8], with *ϵ* = 10%. Even with cloth mask use (*p*_*m*_ = 0.3), these bounds are increased dramatically to 8 and 80 hours, respectively. Assuming six hours of indoor time per day, a school group wearing masks with adequate ventilation would thus be safe for longer than the recovery time for COVID-19 (7-14 days), and school transmissions would be rare. We stress, however, that our predictions are based on the assumption of a “quiet classroom”, where resting respiration (*C*_*q*_ = 30) is the norm. Extended periods of physical activity, collective speech, or singing would lower the time limit by an order of magnitude (Fig. 2).

**Fig. 3.**
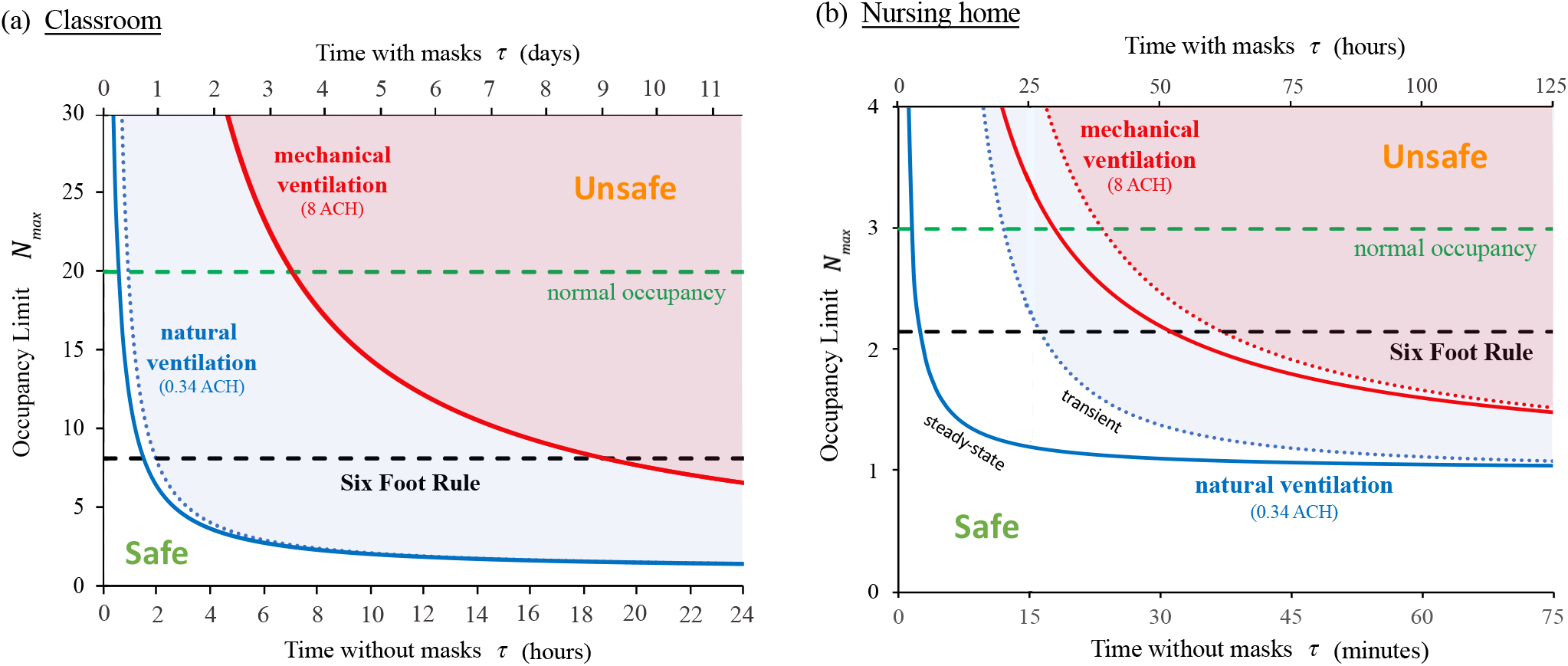
The COVID-19 indoor safety guideline would limit the *cumulative exposure time* in a room with an infected individual to lie beneath the curves shown. Solid curves are deduced from the pseudo-steady formula, Eq. [5], for both natural ventilation (*λ*_*a*_ = 0.34/h; blue curve) and mechanical ventilation (*λ*_*a*_ = 8.0/h; red curve). Horizontal axes denote occupancy times with and without masks. Evidently, the Six-Foot Rule (which limits occupancy to 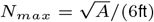) becomes inadequate after a critical time, and the 15-Minute Rule above a critical occupancy. (a) A typical school classroom: 20 persons share a room with an area of 900 square feet and a ceiling height of 12 feet (*A* = 83.6 m^2^, *V* = 301 m^3^). We assume low relative transmissibility (*s*_*r*_ = 25%), cloth masks (*p*_*m*_ = 30%) and moderate risk tolerance (*ϵ* = 10%) suitable for children. (b) A nursing home shared room (*A* = 22.3 m^2^, *V* = 53.5 m^3^) with a maximum occupancy of three elderly persons (*s*_*r*_ = 100%), disposable surgical or hybrid-fabric masks (*p*_*m*_ = 10%) and a low risk tolerance (*ϵ* = 1%) to reflect the vulnerability of the community. The transient formula, Eq. [S8], is shown with dotted curves. Other parameters are *C*_*q*_ = 30 quanta/m^3^, 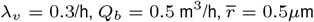.

Our analysis sounds the alarm for elderly homes and long-term care facilities, which account for a large fraction of COVID-19 hospitalizations and deaths (85–87). In nursing homes in New York City, law requires a maximum occupancy of three and recommends a mininum area of 80 square feet per person. In Fig. 3(b), we plot the guideline for a tolerance of *ϵ*= 0.01 transmission probability, chosen to reflect the vulnerability of the community. Once again, the effect of ventilation is striking. For natural ventilation (0.34 ACH), the Six Foot Rule fails after only 3 minutes under quasi-steady conditions, or after 17 minutes for the transient response to the arrival of an infected person, in which case the 15-Minute Rule is only marginally safe. With mechanical ventilation (at 8 ACH) in steady state, three occupants could safely remain in the room for no more than 18 minutes. This example provides insight into the devastating toll of the COVID-19 pandemic on the elderly (85, 87). Furthermore, it underscores the need to minimize the sharing of indoor space, maintain adequate, once-through ventilation, and encourage the use of face masks.

In both examples, the benefit of face masks is immediately apparent, since the CET limit is enhanced by a factor 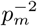, the inverse *square* of the mask penetration factor. Standard surgical masks are characterized by *p*_*m*_ = 1 5% (72, 73), and so allow the cumulative exposure time to be extended by 400– 10000 times. Even cloth face coverings would extend the CET limit by 6–100 times for hybrid fabrics (*p*_*m*_ = 10 − 40%) or 1.5–6 times for single-layer fabrics (*p*_*m*_ = 40 − 80%) (74). Our inference of the efficacy of face masks in mitigating airborne transmission is roughly consistent with studies showing the benefits of mask use on COVID-19 transmission at the scales of both cities and countries (22, 32, 82).

Air filtration has a less dramatic effect than face-mask use in increasing the CET bound. Nevertheless, it does offer a means of mitigating indoor transmission with greater comfort, albeit at greater cost (22, 71). Eq. [5] indicates that even perfect air filtration, *p*_*f*_ = 1, will only have a significant effect in the limit of highly recirculated air, *Z*_*p*_ ≪ 1. The corresponding minimum outdoor airflow per person, *Q/N*_*max*_, should be compared with local standards, such as 3.8 L/s/person for retail spaces and classrooms and 10 L/s/person for gyms and sports facilities (71). In the above classroom example with a typical primary outdoor air fraction of *Z*_*p*_ = 20% (22), the air change rate *λ*_*a*_ could effectively be increased by a factor of 4.6 by installing a MERV-13 filter, *p*_*m*_ = 90%, or by 5.0 with a HEPA filter, *p*_*m*_ = 99.97%. At high air exchange rates, the same factors would multiply the CET bound.

Next, we illustrate the value of our guideline in contact tracing (81), specifically, in prescribing the scope of the testing of people with whom an infected index case has had close contact. The CDC presently defines a COVID-19 “close contact” as any encounter in which an individual is within 6 feet of an infected person for more than 15 minutes. Figure 3 makes clear that this definition may grossly underestimate the number of individuals exposed to a substantial risk of airborne infection in indoor spaces. Our study suggests that whenever our CET bound (5) is violated during an indoor event involving the index case, at least one transmission is likely, with probability *ϵ*. When the tolerance *ϵ* exceeds a critical value, *all* occupants of the room should be considered close contacts and so warrant testing. For relatively short exposures (*λ*_*a*_*τ* ≪ 1) initiated when the index case enters the room, the transient bound should be considered (SI Appendix, Sec. 2).

We proceed by considering the implications of our guideline for the implementation of quarantining and testing. While official quarantine guidelines emphasize the importance of isolating infected persons, our study makes clear the importance of isolating and clearing infected indoor air. In cases of home quarantine of an infected individual with healthy family members, our guideline provides specific recommendations for mitigating indoor airborne transmission. For a group sharing an indoor space intermittently, for example office coworkers or classmates, regular testing should be done with a frequency that ensures that the cumulative exposure time is less than the limit set by the guideline. Such testing would only be unnecessary if the time limit set by the CET bound greatly exceeds the time taken for an infected person to be removed from the population. For the case of a symptomatic infected person, this removal time should correspond to the time taken for the onset of symptoms (≈ 5.5 days). To safeguard against asymptomatic individuals, one should use the recovery time (≈ 14 days) in place of the removal time.

Finally, we briefly discuss the influence of the prevalence of infection in the population on the application of our safety guideline. Our guideline sets a limit on the indoor reproductive number, the risk of transmission from a single infected person in the room. It thus implicitly assumes that the prevalence of infection in the population, *p*_*i*_, is relatively low. In this low-*p*_*i*_ limit, the risk of transmission increases with the expected number of infected persons in the room, *Np*_*i*_, and the tolerance should be lowered in proportion to *Np*_*i*_ if it exceeds one. Conversely, when *Np*_*i*_ → 0, the tolerance might be increased proportionally until the recommended restrictions are deemed unnecessary.

For instructions on how to apply our guideline to other situations, we refer the interested reader to the spreadsheet provided in the SI Appendix. There, by specifying a given room geometry, ventilation rate and respiratory activity, one may deduce the maximum cumulative exposure time in a particular indoor setting, and so define precisely what constitutes an exposure in that setting. An online app based on our guideline has also been developed (101).

## Beyond the Well-Mixed Room

The model developed herein describes the risk of small respiratory drops (*r < r*_*c*_) in the case where the entirety of the room is well mixed. There are undoubtedly circumstances where there are substantial spatial and temporal variations of the pathogen concentration from the mean (7, 41). For example, it is presumably the spatial variations from well-mixedness that result in the inhomogeneous infection patterns reported for a number of well-documented transmission events in closed spaces, including a COVID outbreak in a Chinese restaurant (4), and SARS outbreaks on airliners (102). Circumstances have also been reported where air-conditioner-induced flows appear to have enhanced direct pathogen transport between infected and susceptible individuals (103). In the vicinity of an infected person, the turbulent respiratory jet or puff will have a pathogen concentration that is substantially higher than the ambient (20, 42). Chen *et al*. (41) referred to infection via respiratory plumes as ‘short-range airborne transmission’, and demonstrated that it poses a substantially greater risk than large-drop transmission. In order to distinguish short-range airborne transmission from that considered in our study, we proceed by referring to the latter as ‘long-range airborne’ transmission.

On the basis of the relatively simple geometric form of turbulent jet and puff flows, one may make estimates of the form of the mixing that respiratory outflows induce, the spatial distribution of their pathogen concentration and so the resulting risk they pose to the room’s occupants. For the case of the turbulent jet associated with relatively continuous speaking or breathing, turbulent entrainment of the ambient air leads to the jet radius *r* = *α*_*t*_*x* increasing linearly with distance *x* from the source, where *α*_*t*_ ∼ 0.1 − 0.15 is the typical jet entrainment coefficient (20, 41, 42). The conservation of momentum flux *M* = *π ρ*_*a*_*r*^2^*v*^2^ then indicates that the jet speed decreases with distance from the source according to 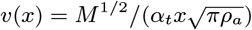. Concurrently, turbulent entrainment results in the pathogen concentration within the jet decreasing according to 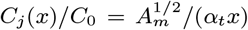, where *A*_*m*_ ∼ 2cm^2^ denotes the cross-sectional area of the mouth and *C*0 = *C*_*q*_ */c*_*v*_ is the exhaled pathogen concentration.^§^ Abkarian *et al*. (42) thus deduce that for the respiratory jet generated by typical speaking, the concentration of pathogen is diminished to approximately 3% of its initial value at a distance of 2 meters.

In a well-mixed room, the mean concentration of pathogen produced by a single infected person is *f*_*d*_*C*_0_. For example, in the large, poorly ventilated room of the Skagit Valley Chorale, we compute a dilution factor, 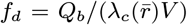 of approximately 0.001. We note that since *λ*_*c*_(*r*) *> λ*_*a*_ = *Q/V*, the dilution factor satisfies the bound, *f*_*d*_ ≤ *Q*_*b*_*/Q*. For typical rooms and air exchange rates, *f*_*d*_ lies in the range of 0.0001 0.01. With the dilution factor of the well-mixed room and the dilution rate of respiratory jets, we may now assess the relative risk to a susceptible person of a close encounter (either episodic or prolonged) with an infected individual’s respiratory jet, and an exposure associated with sharing a room with an infected person for an extended period of time. Since the infected jet concentration *C*_*j*_ (*x*) decreases with distance from its source, one may assess its pathogen concentration relative to that of the well-mixed room, 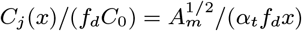. There is thus a critical distance, 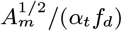 beyond which the pathogen concentration in the jet is reduced to that of the ambient. This distance exceeds 10m for *f*_*d*_ in the afore-mentioned range and so is typically much greater than the characteristic room dimension. Thus, in the absence of masks, respiratory jets may pose a substantially greater risk than the well-mixed ambient.

We first consider a worst-case, close-contact scenario in which a person directly ingests a lung full of air exhaled by an infected person. An equivalent amount of pathogen would be inhaled from the ambient by anyone within the room after a time *τ* = *V*_*b*_*/*(*Q*_*b*_*f*_*d*_), where *V*_*b*_ ≈ 500 mL is the volume per breath. For the geometry of the Skagit choir room, for which *f*_*d*_ = 0.001, the critical time beyond which airborne transmission is a greater risk than this worst-case close encounter with a respiratory plume is *τ* = 1.0 hour. We next consider the worst-case scenario governed by the 6-foot rule, in which a susceptible person is directly in the path of an infected turbulent jet at a distance of 6 feet, over which the jet is diluted by factor of 3% (42). The associated concentration in the jet is still roughly 30 times higher than the steady-state concentration in the well-mixed ambient (when *f*_*d*_ = 0.001), and so would result in a commensurate amplification of the transmission probability. Our guideline could thus be adopted to safeguard against the risk of respiratory jets in a socially-distanced environment by reducing *‘* by a factor of *C*(6*ft*)*/*(*f*_*d*_*C*_0_), which is 3 - 300 for *f*_*d*_ in the range of 0.0001 - 0.01. We note that the latter worst-case scenario describes a static situation where a susceptible individual is seated directly in the respiratory plume of an infected individual, as may arise in a classroom or airplane (102). More generally, with a circulating population in an indoor setting, one would expect to encounter an infected respiratory plume only for some small fraction of the time, consideration of which would allow for a less conservative choice of *ϵ*.

We may thus make a relatively crude estimate for the additional risk of short-range plume transmission, appropriate when masks are not being worn (*p*_*m*_ = 1), by adding a correction to our safety guideline [5]. We denote by *p*_*j*_ the probability that a susceptible neighbor lies in the respiratory plume of the infected person, and by *x >* 0 the distance between nearest neighbours, between which the risk of infection is necessarily greatest. We thus deduce

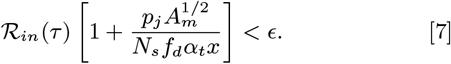

In certain instances, meaningful estimates may be made for both *p*_*j*_ and *x*. For example, if a couple dines at a restaurant, *x* would correspond roughly to the distance across a table, and *p*_*j*_ would correspond to the fraction of the time they face each another. If *N* occupants are arranged randomly in an indoor space, then one expects *p*_*j*_ = tan^*−*1^ *α*_*t*_*/fi* and 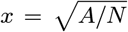. When strict social distancing is imposed, one may further set *x* to the minimum allowed inter-person distance, such as six feet. Substitution from Eq. [5] reveals that the second term in Eq. [7] corresponds to the risk of transmission from respiratory jets deduced by Yang *et al*. (105), aside from the factor *p*_*j*_. We note that any such guideline intended to mitigate against short-range airborne transmission by respiratory plumes will be, as is [7], dependent on geometry, flow and human behavior, while our guideline for the mitigation of long-range airborne transmission [5] is universal.

We note that the use of face masks will have a marked effect on respiratory jets, with the fluxes of both exhaled pathogen and momentum being reduced substantially at their source. Indeed, Chen *et al*. (41) note that when masks are worn, the primary respiratory flow may be described in terms of a rising thermal plume, which is of significantly less risk to neighbours. With a population of individuals wearing face masks, the risk posed by respiratory jets will thus be largely eliminated, while that of the well-mixed ambient will remain.

Finally, we stress that our guideline is based on the *average* concentration of aerosols within the room. For every region of enhanced airborne pathogen concentration, there is necessarily a region of reduced concentration and lower transmission risk elsewhere in the room. The ensemble average of the transmission risk over a number of similar events, and the time-averaged transmission risk in a single event are both expected to approach that in the well-mixed steady state, as in ergodic processes in statistical mechanics. This feature of the system provides rationale for the self-consistency of our inferences of *C*_*q*_, based on the hypothesis of the well-mixed room, from the diverse set of spreading events considered herein.

## Discussion and Caveats

We have focused here primarily on airborne transmission, for which infection arises through inhalation of a critical quantity of airborne pathogen, and neglected the roles of both contact and large-drop transmission (6). While motivated by the COVID-19 pandemic, our theoretical framework applies quite generally to airborne respiratory illnesses, including influenza. Moreover, we note that the approach taken, coupling the droplet dynamics to the transmission dynamics, allows for a more complete description. For example, consideration of conservation of pathogen allows one to calculate the rate of pathogen sedimentation and associated surface contamination, consideration of which would allow for quantitative models of contact transmission and so inform cleaning protocols.

Typical values for the parameters arising in our model are listed in SI Appendix, Table S1. Respiration rates *Q*_*b*_ have been measured to be ∼0.5m^3^/hr for normal breathing, and may increase by a factor of 3 for more strenuous activities (17). Other parameters, including room geometry, ventilation and filtration rates, will obviously be room dependent. The most poorly constrained parameter appearing in our guideline is *C*_*q*_*s*_*r*_, the product of the concentration of pathogen in the breath of an infected person and the relative transmissibility. The latter, *s*_*r*_, was introduced in order to account for the dependence of transmissibility on the mean age of the population (85–87, 90) and the viral strain (83, 84). The value of *C*_*q*_*s*_*r*_ was inferred from the the best characterized super-spreading event, the Skagit Valley Chorale incident (24), as arose amongst an elderly population with a median age of 69 (26), for which we assign *s*_*r*_ = 1. The *C*_*q*_ value so inferred was rescaled using reported drop-size distributions (9, 23, 37) allowing us to estimate *C*_*q*_ for several respiratory activities, as listed in Figure 3. Further comparison with inferences based on other spreading events of new viral strains among different populations would allow for refinement of our estimates of *C*_*q*_ and *s*_*r*_. We thus appeal to the public health community to document the physical conditions enumerated in Table S1 of SI Appendix for more indoor spreading events.

Adherence to the Six-Foot Rule would limit large-drop transmission, and to our guideline, Eq. [5], would limit long-range airborne transmission. We have also shown how the sizeable variations in pathogen concentration associated with respiratory flows, arising in a population not wearing face masks, might be taken into account. Consideration of both short-range and long-range airborne transmission leads to a guideline of the form of Eq. [7] that would bound both the distance between occupants and the cumulative exposure time. Circumstances may also arise where a room is only partially mixed, owing to the absence or deficiency of air conditioning and ventilation flows, or the influence of irregularities in the room geometry (106). For example, in a poorly ventilated space, contaminated warm air may develop beneath the ceiling, leading to the slow descent of a front between relatively clean and contaminated air a process described by ‘filling-box’ models (106). In the context of reducing COVID-19 transmission in indoor spaces, such variations from well-mixedness need be assessed on a room-by-room basis. Nevertheless, the criterion [5] represents a minimal requirement for safety from long-range airborne infection in well-mixed, indoor spaces.

We emphasize that our guideline was developed specifically with a view to mitigating the risk of long-range airborne transmission. We note, however, that our inferences of *C*_*q*_ came from a number of super-spreading events, where other modes of transmission, such as respiratory jets, are also likely to have contributed. Thus, our estimates for *C*_*q*_ are necessarily *over-estimates*, expected to be higher than those that would have arisen from purely long-range airborne transmission. Consequently, our safety guideline for airborne transmission necessarily provides a conservative upper bound on cumulative exposure time. We note that the additional bounds required to mitigate other transmission modes will not be universal; for example, we see in Eq. [7] that the danger of respiratory jets will depend explicitly on the arrangement of the room’s occupants. Finally, we reiterate that the wearing of masks largely eliminates the risk of respiratory jets, and so makes the well-mixed-room approximation considered here all the more relevant.

Our theoretical model of the well-mixed room was developed specifically to describe airborne transmission between a fixed number of individuals in a single well-mixed room. Nevertheless, we note that it is likely to inform a broader class of transmission events. For example, there are situations where forced ventilation mixes air between rooms, in which case the compound room becomes effectively a well-mixed space. Examples considered here are the outbreaks on the Diamond Princess and in apartments in Wuhan City (see SI Appendix); others would include prisons. There are many other settings, including classrooms and factories, where people come and go, interacting intermittently with the space, with infected people exhaling into it, and susceptible people inhaling from it, for limited periods. Such settings are also informed by our model provided one considers the mean population dynamics, specifically, that *N* be identified as the mean number of occupants.

The guideline [5] depends on the tolerance *‘*, whose value in a particular setting should be set by the appropriate policy makers, informed by the latest epidemiological evidence. Likewise, the guideline includes the relative transmissibility *s*_*r*_ of a given viral strain within a particular sub-population. These two factors may be eliminated from consideration by using to assess the *relative behavioral risk* posed to a particular individual by attending a specific event of duration *τ* with *N* other participants. We thus define a risk index,

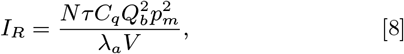

that may be evaluated using appropriate *C*_*q*_ and *Q*_*b*_ values (listed in SI Appendix, Fig. S2). One’s risk increases linearly with the number of people in a room and duration of the event. Relative risk decreases for large, well ventilated rooms and increases when the room’s occupants are exerting themselves or speaking loudly. While these results are intuitive, the approach taken here provides a physical framework for understanding them quantitatively. It also provides a quantitative measure of the relative risk of certain environments, for example, a well ventilated, sparsely occupied laboratory and a poorly ventilated, crowded, noisy bar. Along similar lines, the weighted average of (8), provides a quantitative assessment of one’s risk of airborne infection over an extended period. It thus allows for a quantitative assessment of what constitutes an *exposure*, a valuable notion in defining the scope of contact tracing, testing and quarantining.

Above all, our study makes clear the inadequacy of the Six-Foot Rule in mitigating indoor airborne disease transmission, and offers a rational, physically-informed alternative for managing life in the time of COVID-19. If implemented, our safety guideline would impose a limit on the cumulative exposure time in indoor settings, violation of which constitutes an exposure for all of the room’s occupants. Finally, while our study has allowed for an estimate of the infectiousness of COVID-19, it also indicates how new data characterizing indoor spreading events may lead to improved estimates thereof and so to quantitative refinements of our safety guideline.

The spreadsheet included in Dataset S1 provides a simple means of evaluating this limit for any particular indoor setting. A convenient online app based on our safety guideline is also available (101). A glossary of terms arising in our study is presented in Table S3 of the SI Appendix.

## Supporting information

COVID-19 Indoor Safety Guideline Calculator

## Data Availability

An Excel spreadsheet is provided as a supplementary file to calculate our COVID-19 safety guideline for a specific indoor space. Expiratory aerosol droplet size distributions were taken from the cited literature, as either provided by the authors (Asadi et al, 2020) or digitally scanned from published figures (Morawska et al, 2009).

http://www.mit.edu/~bazant/COVID-19

https://indoor-covid-safety.herokuapp.com

## ACKNOWLEDGMENTS

The authors thank William Ristenpart and Sima Asadi for sharing experimental data and Lesley Bazant, Lydia Bourouiba, Daniel Cogswell, Mark Hampden-Smith, Kyle Hofmann, David Keating, Lidia Morawska, Nels Olson, Monona Rossol, and Renyi Zhang for important references.

## Footnotes

* The possibility of pathogen resuspension from contaminated surfaces has also recently been explored (10, 11).

† For the sake of simplicity, we do not consider here the dependence of *p*_*m*_ on respiratory activity (76) or direction of airflow (77), but note that, once reliably characterized, these dependencies might be included in a straightforward fashion.

‡ Markov’s inequality ensures that the probability of at least one transmission, *P*_1_, is bounded above by the expected number of transmissions, *P*_1_ ≤ *R*_*in*_. In the limit, *R*_*in*_ *< ϵ ≪* 1, these quantities are asymptotically equal, since 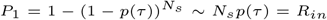 for *N*_*s*_ independent transmissions of probability, 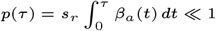.

§ These expressions for *v*(*x*) and *C*(*x*) are valid in the limit of *x > x*_*v*_ where *x*_*v*_ is the virtual origin of the jet, typically on the order of 10cm (20, 104). Near-field expressions well behaved at *x* = 0 are given by replacing *x* with *x* + *x*_*v*_, and normalizing such that *C*(0) = *C*_0_.

