## Supplementary Information (Appendices) for "Beyond Six Feet: A Guideline to Limit Indoor Airborne Transmission of COVID-19"

### 1. Validity of the model assumptions

We proceed by providing rationale for the two key assumptions of our model, and discussing their range of validity.

**A. The well-mixed room.** Two flow types typically contribute to vigorous mixing of indoor air: buoyancy-driven and forced convection. Buoyancy-driven convection results from the action of gravity on differences in air density, typically due to temperature gradients. Forced-air heating (from below) or cooling (from above) of a room typically lead to localized flows that are subject to hydrodynamic instabilities that prompt mixing on a relatively larger scale. Air-conditioning units generate relatively cold, dense air that sinks as a turbulent plume and entrains ambient air as it sinks to the floor (1). Heating vents and radiators create buoyant counterparts, turbulent thermal plumes that rise to the ceiling. In relatively cool rooms, temperature gradients associated with body heat lead to turbulent thermal plumes rising from individuals. Respiratory jets, plumes and puffs also typically have some buoyancy, leading to updrafts from these turbulent respiratory flows (2). When masks are worn, the horizontal momentum of respiratory flows is greatly suppressed, but the exhaled air typically leads to a rising turbulent buoyant plume (3). Additional large-scale flows, either laminar or turbulent, are driven by horizontal temperature gradients in the vicinity of closed windows. The net effect of such buoyancy-driven flows is to promote mixing of the indoor air.

Room-scale forced convection may also be driven by the mechanical air circulation system or the respiration and movement of its occupants. The hydrodynamic stability of an air flow with a characteristic speed $v$ and length scale $L$ is prescribed by the Reynolds number, $\mathrm{Re} = vL/\nu_a$, where $\nu_a$ is the kinematic viscosity of air. For example, laminar flow around obstacles typically becomes unstable to vortex shedding for $\mathrm{Re} > 100$ and turbulence for $\mathrm{Re} > 2000$. For an indoor air flow forced by mechanical ventilation with a characteristic speed $v = (Q + Q_r)/A = (\lambda_a + \lambda_r)H$ and length scale corresponding to that of a room, $L = H = 3\mathrm{m}$, the characteristic Reynolds number $\mathrm{Re} \approx 110$ for $\lambda_a = 0.3/\mathrm{h}$ and $\mathrm{Re} \approx 2000$ for $\lambda_a = 8/\mathrm{h}$, even in the absence of recirculation flow ($\lambda_r = 0$). Thus, forced convection will typically lead to high-Reynolds-number flows that may be characterized by some combination of vortex shedding and turbulence. Use of fans, air-filtration units or open windows will further increase airflow and mixing. Human movement may also generate vigorous, high-Reynolds-number flows. Human respiration without masks leads to turbulent puff trains and jets (4), coughing and sneezing to turbulent, buoyant puff clouds (2).

In turbulent flows, the largest eddies are at the scale of the confining geometry and control the convective transport. Turbulent mixing by forced convection at the scale of the room is thus characterized by the eddy diffusivity, $D_e \approx \frac{1}{2}vH$, as has been verified for the transport of tracer gas in instrumented homes (5). The characteristic mixing time, $\tau_{mix} = H^2/2D_e = H/v = (\lambda_a^{-1} + \lambda_r^{-1})^{-1}$, is thus prescribed by the time scale of the faster of the two processes, outdoor air exchange and internal air recirculation. In either case, the room should be well mixed by the time the bioaerosol concentration reaches its steady state, since the concentration relaxation rate necessarily exceeds the air exchange rate due the influence of droplet settling, filtration and viral deactivation.

In summary, indoor air is typically well mixed by either laminar or turbulent flows driven by some combination of mechanical air circulation, buoyancy-driven flows and human activity (6). The success of turbulent deposition models (7–10) in rationalizing measured aerosol mass transfer to indoor surfaces (7, 11, 12) provides further support for the validity of the well-mixed-room approximation. While variations from well-mixedness may arise, for example through the development of stratification (6), the approximation of the well-mixed room has been widely applied in describing indoor settings, specifically in the context of long-range airborne disease transmission (13–17).

**B. Droplet transfer to surfaces .** In the continuous stirred tank reactor (CSTR)(18), a canonical system in chemical engineering, the relative rate of chemical reaction to convection is prescribed by the dimensionless Damköller number. In the context of airborne disease transmission, we consider the 'reaction' corresponding to aerosol-borne virion removal from a well-mixed room, and so define $\mathrm{Da} = \lambda_c/\lambda_a$ in terms of the concentration relaxation rate $\lambda_c(r)$ and the air change rate $\lambda_a$. To distinguish between the distinct contributions to this

[1] To whom correspondence should be addressed. E-mail: bazantmit.edu

reaction from bulk and surface removal processes, we write

$$\mathrm{Da} = \mathrm{Da}_b + \mathrm{Da}_s = \frac{(\lambda_v + \lambda_f)V}{Q} + \frac{hA}{Q}\ . \qquad \text{[S1]}$$

Bulk disinfection of the air may arise through viral deactivation at a rate $\lambda_v(r)$ or air filtration at a rate $\lambda_f(r) = p_f(r)\lambda_r$. We denote by $h$ the virion mass-transfer coefficient from the bulk to the surfaces, which necessarily has the units of velocity.

In the field of aerosol science (7, 8, 19), it is customary to refer to the dimensionless droplet mass-transfer coefficient as the 'deposition efficiency', $\eta = hA/Q$, and to relate it to various particle transport resistances. Such resistances are expressed in dimensionless form in terms Stokes numbers (12), each of which represents the ratio of two particle speeds. There are three potentially relevant resistances to drop mass-transfer from the bulk air to surfaces: gravitational settling, diffusion and surface deposition. We thus express the efficiency of droplet deposition from the bulk in terms of three Stokes numbers:

$$\eta^{-1} = (\mathrm{St}_g + \mathrm{St}_d)^{-1} + \mathrm{St}_{dep}^{-1}. \qquad \text{[S2]}$$

We note that the droplet deposition efficiency $\eta$ is equivalent to the "particle loss-rate coefficient", $\beta$, customarily defined for particulate aerosols in indoor air (7))

The gravitational Stokes number, $St_g = v_s/v_a$, prescribes the relative magnitudes of the Stokes settling speed, $v_s = 2\rho_d g r^2/9\mu_a$ and the ambient air flow, $v_a = Q/A = \lambda_a L$. We note that inertial corrections to Stokes law (20) only become important for droplets with $r > 100\mu$m, and so are not relevant for aerosols. For the largest respirable droplets ($r = 5\mu$m) with poor, natural ventilation (0.35 ACH) in a typical room ($H = 2.7$m), settling dominates convection since $\mathrm{St}_g = 12 \gg 1$. Conversely, for smaller droplets ($r = 0.5\mu$m) with hospital-grade ventilation (25 ACH), settling can be neglected since $\mathrm{St}_g = 0.0017 \ll 1$. Drops with radius exceeding a critical values, $r_c = \sqrt{9\mu_a\lambda_a H/(2\rho_d g)}$ corresponding to $\mathrm{St}_g = 1$, tend to settle to the surfaces before being removed by ventilation. For a given droplet size $r$, we write $\mathrm{St}_g(r) = (r/r_c)^2$. The droplets of interest in our model of airborne transmission are those with $St_g < 1$.

For submicron particles suspended in a well-mixed ambient, diffusion may influence the transfer of the particle through the lower viscous boundary layer, either by Brownian motion or eddy diffusion for the case of a turbulent boundary layer (7, 12, 19). The importance of such particle diffusion is prescribed by the diffusional Stokes number (12), $St_d = D/(\delta v_a)$, where $D$ is the particle diffusivity and $\delta$ is the viscous boundary layer thickness near the depositing surface, which sets the diffusional mass-transfer coefficient, $h_d = D/\delta$. We may alternatively write $St_d = \overline{\mathrm{Sh}}(\mathrm{Re}, \mathrm{Sc})/\mathrm{Pe}$, where the Péclet number, $\mathrm{Pe} = v_a H/D$, prescribes the relative magnitudes of convection and diffusion, and the Sherwood number, $\overline{\mathrm{Sh}} = h_d H/D$, is the dimensionless diffusion mass-transfer coefficient, averaged over the surface.

We first consider Brownian droplet diffusion through a laminar boundary layer, as is typical of natural ventilation over horizontal surfaces. For a given flow field, kinematic viscosity $\nu_a = \mu/\rho_a$ and Brownian diffusivity $D = k_B T/6\pi\mu_a r_d$ (as follows from the Stokes-Einstein relation), the Sherwood number depends on the Schmidt number $\mathrm{Sc} = \nu_a/D$ and Reynolds number $\mathrm{Re} = \mathrm{Pe}/\mathrm{Sc}$. Using the relation, $\overline{\mathrm{Sh}} = 0.664\,\mathrm{Re}^{1/2}\mathrm{Sc}^{1/3}$, relevant for a laminar boundary layer on a flat plate (21), we estimate, for the case of bioaerosols with radius ($r = 0.5\mu$m) with poor ventilation (0.35 ACH), that $\mathrm{St}_d = 7 \times 10^{-6} \ll 1$ ($\mathrm{Sc} = 5 \times 10^5$, $\mathrm{Re} = 47$, $\mathrm{Sh} = 180$, $\mathrm{Pe} = 2.5 \times 10^7$), which indicates that particle diffusion can be safely neglected. Indeed, the removal of droplets by Brownian diffusion becomes comparable to that by gravitational settling, $\mathrm{St}_d > \mathrm{St}_g$, only for $r < 13$ nm at 0.35 ACH, a critical radius much smaller than that of a single SARS-CoV-2 virion ($r_v = 60$nm).

Next, we consider droplet diffusion through a turbulent boundary layer, as might arise for high ventilation rates. In their seminar work on aerosol mass transfer, Corner and Pendleton (8) augmented the Brownian diffusivity, $D$, with the turbulent eddy diffusivity, $D_e = K_e y^2$, where $K_e$ is a turbulence intensity parameter, proportional to the local mean velocity gradient, and $y$ is the distance from the surface. A better fit to experimental data was achieved by expressing the eddy diffusion as $D_e = K_e\delta^2(y/\delta)^n$ (9, 10), where $\delta$ is the boundary layer thickness $\delta$ and $n = 2.6 - 2.8$ is an empirical scaling exponent (7). Using this approach, Lai & Nazaroff (7) developed a comprehensive, experimentally validated model of aerosol mass transfer to surfaces and showed that, even for the highest building ventilation rates, turbulent diffusion competes with gravitational settling only for particles with $r < 0.2\mu$m. This critical size is much smaller than that of stable respiratory droplets (22–24), and only a few times larger than a single virion. We thus conclude that gravitational settling dominates diffusion, both Brownian and turbulent, in respiratory aerosol removal from indoor air.

Finally, the role of droplet deposition is determined by $\mathrm{St}_{dep} = v_{dep}/v_a = \mathrm{St}_g v_{dep}/v_s$, where $v_{dep}$ is the net liquid deposition velocity, specifically the surface adsorption flux per bulk liquid concentration, as accounts for the probability of droplets sticking to the surface rather than bouncing off. Experiments have shown that $v_{dep}/v_s$ is approximately constant for water droplets with characteristic diameter of 1-10 $\mu$m in a variety of settings, including droplets settling from clouds onto the forest canopy(25) and from humid air onto acrylic pipes(26, 27)). For droplets of radius $r = 5\mu$m settling at $v_s = 0.306$ cm/s, the deposition velocity $v_{dep} \approx 5$ cm/s. We thus conclude that deposition kinetics are fast compared to droplet transport, since $\mathrm{St}_{dep} \approx 20\,\mathrm{St}_g$, and can be safely neglected as a small mass-transfer resistance in series.

In summary, for the respiratory droplets of interest, the dominant contribution to droplet mass transfer to surfaces for the relevant range of respiratory aerosols, $r = 0.1 - 10\mu m$ comes from gravitational settling, $h \approx v_s$. We are thus justified in writing

$$\eta \approx \mathrm{St}_g = \left(\frac{r}{r_c}\right)^2 \qquad \text{[S3]}$$

as assumed in the main text. This deduction allows us to conveniently express the droplet mass transfer from complex indoor airflows in terms of the Stokes settling velocity. The effects of boundary-layer or surface mass-transfer resistance are negligible, and the timescale of settling of respiratory droplets from a well-mixed ambient is prescribed by the Stokes settling time.

### 2. Exact solution for the transient guideline

The general safety guideline, Eq. [3], can be written as, $\mathcal{R}_{in}(\tau) = N_s s_r \langle\beta_a\rangle\tau$, in terms of the time-averaged airborne transmission rate, which can be broken into steady-state and transient terms:

$$\langle\beta_a\rangle(\tau) = \frac{1}{\tau}\int_0^\tau \beta_a(t)dt = \overline{\beta}_a - \Delta\beta_a(\tau) \quad \text{[S4]}$$

$$\overline{\beta}_a = \frac{Q_b^2}{V}\int_0^\infty \frac{n_q(r)}{\lambda_c(r)}dr \quad \text{[S5]}$$

$$\Delta\beta_a(\tau) = \frac{Q_b^2}{V\tau}\int_0^\infty \frac{n_q(r)}{\lambda_c(r)^2}\left(1-e^{-\lambda_c(r)\tau}\right)dr \quad \text{[S6]}$$

where $n_q(r) = n_d(r)V_d(r)c_v(r)c_i(r)$ and $\lambda_c(r) = \lambda_a(1 + (r/r_c)^2) + \lambda_v(r) + \lambda_f(r)$. The steady-state term $\overline{\beta}_a$ can be expressed as Eq. [4], where the mean sedimentation speed $v_s(\bar{r})$ and mean suspended droplet size, $\bar{r}$, are defined by

$$\frac{v_s(\bar{r})}{\nu(\bar{r})^2 v} = \left(\frac{\bar{r}}{\nu(\bar{r})r_c}\right)^2 = C_q\left(\int_0^\infty \frac{n_q(r)dr}{1+(r/(\nu(r)r_c))^2}\right)^{-1} - 1 \quad \text{[S7]}$$

with $v = Q/A$ and $\nu(r) = \sqrt{1 + (\lambda_v(r) + \lambda_f(r))/\lambda_a}$, which gives most weight in the average to the suspended aerosol droplets with $r < r_c$.

For short exposures or poor ventilation, the transient correction can reduce the indoor reproductive number, resulting in a more permissive guideline. In the case of monodisperse droplets of size $r = \bar{r}$, the transient term can be approximated as, $\Delta\beta_a/\overline{\beta}_a = (1 - e^{-\lambda_c\tau})/(\lambda_c\tau) \approx 1/(1 + \lambda_c\tau)$. We may thus recast the general safety guideline in the form,

$$N_s s_r \overline{\beta}\tau < \epsilon(1 + (\lambda_c(r)\tau)^{-1}) \quad \text{[S8]}$$

which reduces to the pseudo-steady criterion, $N_s s_r \tau\overline{\beta} < \epsilon$ for exposure times long relative to the concentration relaxation time, $\lambda_c(r)\tau \gg 1$. In the limit of short exposures, $\lambda_c(r)\tau \ll 1$, we obtain a refined safety guideline, $N_s s_r(\overline{\beta}\tau)(\lambda_c(r)\tau) < \epsilon$, which is less strict because it reflects the leeway associated with the time taken for the build-up of the airborne pathogen following the arrival of an infected person. During this period, the safe exposure time scales as $\tau_{max} \sim N_s^{-1/2}$, and the CET bound diverges as $N_s\tau_{max} \sim \tau^{-1}$ in the $\tau \to 0$ limit. This divergence simply reflects the fact that transmission will not occur if people do not spend sufficient time together.

### 3. Deduction of the indoor safety guideline from epidemiological models

We here demonstrate that our indoor safety guideline, Eq. [3], may be deduced from standard epidemiological models.

While our theoretical model of the well-mixed room was developed to describe airborne transmission from a single individual to a fixed number of others in a well-mixed room, it bears noting that this scenario is but one of a broader class of transmission events. First, there are many situations where air conditioning or forced ventilation mixes air between rooms, in which case the compound room is effectively a well-mixed space. Second, there are incidents when several individuals are initially infected. Third, the population may be together for a period long with respect to the incubation time (approximately 5.5 days (28)), in which case the number of infectious individuals necessarily increases with time. Examples of this broader class of transmission events might include cruise ships (29–33), meat and poultry processing facilities (34, 35) and prisons (36, 37). We proceed by detailing the epidemiological models describing this more general class of events.

Following Noakes et al.(38), we adopt the standard SEIR model from epidemiology (39–41), an extension of the original SIR model of Kermack and McKendrick(42) of susceptible ($S$), infected ($I$) and recovered ($R$) populations to include an exposed population ($E$) that has acquired the pathogen and so become infected but not yet infectious:

$$\frac{dS}{dt} = -\beta SI,\ \frac{dE}{dt} = \beta SI - \alpha E,\ \frac{dI}{dt} = \alpha E - \gamma I,\ \frac{dR}{dt} = \gamma I \quad \text{[S9]}$$

where $\alpha$ is the incubation rate, $\beta(t)$ is the transmission rate (setting $s_r = 1$), and $\gamma$ is the recovery rate of the infected. To describe a spreading event in a closed environment, we consider $I_0$ infected people entering, at time $t = 0$, a room full of $N_s = S_0 = N - I_0$ susceptible individuals (setting $p_s = 1$), none of which were previously exposed, $E_0 = 0$. The reproductive number, $\mathcal{R}_0 = S_0\beta/\gamma$, describes the initial exponential rate of increase ($\mathcal{R}_0 > 1$) or decrease ($\mathcal{R}_0 < 1$) of the infected fraction of the population (39). In the case where the population is being tested and infected individuals removed from the population via isolation, $\gamma$ may alternatively be considered as a proxy for testing frequency. Noting that the combined incubation time and time to recovery for COVID-19 is thought to exceed 2 weeks (43), we here consider spreading events of duration $\tau$ that are short relative to the recovery time, $\tau \ll \gamma^{-1}$, and so eliminate $R$ from consideration.

The Wells-Riley model is effectively a reduction of the SIR model based on the assumption of slow incubation over the timescale of the event, $\tau \ll \alpha^{-1}$, in which case the number of infected persons in the room remains fixed, $I(t) = I_0$. One expects such to be the case for events of relatively short duration, such as the Skagit Choir incident. However, such an approximation is not likely to be valid in the case of indoor super-spreading events of longer duration, $\tau > \alpha^{-1}$. To adequately model such events, we consider the possibility that the number of infectors increases with time, which requires a description of the nonlinear dynamics of disease transmission.

The resulting SEI model to be considered here is a system of three nonlinear ordinary differential equations. While the system may be solved numerically, deduction of a safety guideline requires analytical results. Our goal is thus to solve this SEI model for the infection (or "secondary attack") rate, $i_S(\tau) = (E(\tau) + I(\tau) - I_0)/S_0$. Since exact solutions may be cumbersome (as for the SIR model (44)), we simplify the analysis by considering two limits, where the incubation time $\alpha^{-1}$ is short or long relative to the event timescale, $\tau$, and the transmission timescale $\beta^{-1}$. While the incubation time of COVID-19 is not precisely known, it is bounded above by the time to develop symptoms, which spans 2-14 days with an average 5.5 days (28).

For slow incubation ($\alpha \ll \beta, \tau^{-1}$) (38), the number of infected persons remains nearly constant, $I(t) \approx I_0$, and the resulting SE model is easily solved to derive the secondary attack rate (45),

$$i_S(\tau) = 1 - e^{-I_0\int_0^\tau \beta(t)dt}, \quad \text{[S10]}$$

In the opposite limit of fast incubation ($\alpha \gg \beta, \tau^{-1}$), exposed individuals are rapidly infected, so the exposed, non-infected population need not be considered, $E(t) = 0$. Solving the resulting SI model yields

$$i_S(\tau) = \left[1 - i_0^{-1}\left(e^{N\int_0^\tau \beta(t)dt} - 1\right)^{-1}\right]^{-1}, \qquad \text{[S11]}$$

where $i_0 = I_0/N$ is the initial percentage of infected individuals. These simple solutions, (S10) and (S11), cover the two limiting cases of fast and slow incubation relative to transmission.

The safety criterion $\mathcal{R}_{in} < \epsilon$, which limits the probability of the first transmission to lie below a small tolerance, is universal, in that it does not depend on the choice of model to describe the subsequent spreading dynamics. In particular, the safety criterion, $I(\tau) < 1 + \epsilon$, with $I_0 = 1$ corresponds to

$$\frac{\mathcal{R}_{in}}{N-1} = \int_0^\tau \beta(t)dt < \begin{cases} -\ln\left(1 - \frac{\epsilon}{N-1}\right) & \text{(slow)} \\ \frac{1}{N}\ln\left[\frac{(N-1)(1+\epsilon)}{N-1-\epsilon}\right] & \text{(fast)} \end{cases} \qquad \text{[S12]}$$

according to Eqs. (S10) and (S11) for slow and fast incubation, respectively. In the small-tolerance limit, $\epsilon \ll 1$, these bounds both reduce to, $\mathcal{R}_{in} < \epsilon$, and are thus independent of the incubation rate. More generally, the linear response of any Markovian, mean-field theory (with neither memory nor many-body correlations) to any perturbation, such as the arrival of a new infector, must begin at the mean transmission rate. Indeed, the same universal bound may be simply derived from the infinitesimal cumulative probability of transmission $\int_0^\tau \beta(t)$ from one infector to a set of $N-1$ independent susceptibles.

### 4. Inference of infection quanta from disease spreading data

It is an important point that the concentration of "infection quanta" per exhaled air volume, $C_q$, as may be inferred from spreading data, is necessarily model dependent. By definition, the time integral of the transmission rate, $p(\tau) = \int_0^\tau \beta(t)dt$, is equal to the expected number of quanta transferred from one infector to one susceptible. In models of indoor airborne transmission (16, 38, 45), infection quanta are further related to the evolving pathogen-laden droplet concentration by Eq. [4]. While the mathematical definition of infection quanta is unambiguous in terms of the instantaneous transmission rate, $\beta(t)$, between two isolated individuals, the inference of quanta concentrations from field data for the spreading rate is inevitably model dependent because it involves interactions among an evolving population. Specifically, the long-time growth in the number of infections is influenced by nonlinearities in the transmission dynamics, as governed by the chosen epidemiological model, for example, the SEIR model, Eq. [S1].

In the original Wells-Riley model (38, 45–48), one infection quantum is defined as the amount of pathogen required to infect an average of $1 - 1/e \approx 63.2\%$ of susceptible people in an enclosed space. This inference is based on fitting field data to Eq. [S2], which relates one net quantum transferred, $I_0 q(t) = 1$, to the secondary infection rate, $i_S(t) = 1 - (1/e)$. This approach is valid for times short relative to the incubation time and has been successfully applied to extract quanta emission rates for various viral diseases (49–52), including SARS-CoV-2 (16, 53). However, the Wells-Riley model cannot be reliably used to infer infection quanta from super-spreading events evolving over time scales that exceed the incubation time.

For such situations, here we take an approach appropriate for the case of fast incubation that necessarily implies a different relation between released infection quanta and the observed secondary infection rate. If we adopt the SI model with fast incubation, then the long-term behavior of Eq. [S3] indicates that one quantum ($I_0 q(t) = 1$) will infect a fraction $i_S = [1 + (N/I_0)/(e^{N/I_0} - 1)]^{-1}$ of $N$ people in an enclosed space, $I_0$ of which were initially infected, as a result of transmission amplification by the growing number of infectors. In this model, a single quantum from one person infects another with probability $i_S = [1 + 2/(e^2 - 1)]^{-1} \approx 76.2\%$ for $N = 2$, but manages to infect everyone in a large group, $i_S \to 1$ as $N \to \infty$. This dependence of infection quanta on epidemiological model parameters, such as the incubation rate, reflects the fact that the fitted "infection quantum" is a measure of contagiousness at the scale of a group that is not necessarily proportional to the microscopic pathogen concentration. Notably, infection quanta are well defined in the limit of short times and slow transmission, where all models reduce to the Wells-Riley model with a constant number of infectors. From the modeling perspective, the notion of infection quanta is thus unambiguous only in this limit. Finally, we recall that, regardless of the model used to infer it, the actual value of infection quanta may still vary considerably between spreading events, owing to its dependence on activity level of the population and other physiological factors (16, 54).

### 5. Fitting to super-spreading events

Table S1 shows the data used to infer the concentration of infection quanta $C_q$ exhaled by an infected individual for four well-known indoor super-spreading events of COVID-19, for which physical parameters can be estimated with reasonable accuracy from published data: the Skagit Valley Chorale (43, 53), the Diamond Princess cruise ship (30, 31), the Ningbo tour bus (55, 56), and the initial Wuhan outbreak (64), interpreted as mainly indoor spreading at home (60). The best characterized case of the Skagit Valley Chorale is used in the main text to calibrate the safety guideline. Here we consider the other three spreading events, for which only relatively rough estimates of certain model parameters can be made. Despite the resulting uncertainty, these events provide additional support for the inferences made in the main text.

In all situations, we assume an effective particle radius $\bar{r} = 2.0\mu$m at the upper limit of the infectious range of suspended droplets (65), corresponding to an effective settling speed of $v_s(\bar{r}) = 1.78$ m/h. Note that the effective radius would be somewhat larger, $\bar{r} = 3.0\mu$m, if we were to neglect the size dependence of infectivity and set $n_q =$constant, and use our prediction of the steady-state airborne droplet distribution $C_s(r)/c_v$ for singing in the choir room (Fig. 1). We also assume no use of face masks, $p_m = 1$ and an airborne virus deactivation rate, $\lambda_v = 0.3$/h (53) that lies between the existing estimates of zero (66) and 0.63/h (67)

**A. The Diamond Princess.** The data reported for the Diamond Princess is particularly useful in that it captures the time evolution of the infected persons among a fixed population over an extended period, corresponding to the 12-day quarantine (30, 31), after which passengers and crew began to disembark. The resulting data reported for $I(t)$ is best described in terms of the fast-incubation limit (Figure S1). Fit-

| | $N$ | $\tau$ (h) | $I_0$ | $I(\tau)$ | $\lambda_a$ (1/h) | $A$ ($m^2$) | $H$ (m) | $Q_b$ ($m^3$/h) | $C_q$ (q/$m^3$) | $\lambda_q$ (q/h) |
|---|---|---|---|---|---|---|---|---|---|---|
| Skagit Church Choir | 61 | 2.5 | 1 | 53 | 0.65 | 180 | 4.5 | 1.0 | 870 | 870 |
| Ningbo Tour Bus | 68 | 1.7 | 1 | 21 | 1.25 | 25 | 1.8 | 0.5 | 90 | 45 |
| Diamond Princess | 3711 | 288 | 20 | 354 | 8 | 139,000 | 2.1 | 0.5 | 30 | 15 |
| Wuhan City Outbreak | 3.03 | 132 | 1 | 1.63 | 0.34 | 90 | 2.4 | 0.5 | 29 | 14 |

**Table S1. Data from four COVID-19 spreading events used to infer the concentration, $C_q$, or emission rate, $\lambda_q$, of exhaled infection quanta, on the basis of the assumption of indoor airborne transmission. 1. The Skagit Valley Choir event (43). We use existing estimates of relevant physical parameters (53) and the Wells-Riley model (16, 45–47, 53), Eq. [S10], appropriate for slow incubation. 2. A tour bus transported 68 people (including the driver) on a 100 minute round-trip journey to a Buddhist ceremony in Ningbo, China (55, 56). One index case infected 23 fellow passengers, three of which are assumed to have been infected at the ceremony, where the infection rate was $7/172 = 4\%$ for other attendees. The interior bus dimensions are estimated from a Dongfeng 67-seat luxury tour bus made in Hubei, China, which has outer dimensions of 10.49m×2.5m×3.2m and matches the floor plan of the Ningbo tour bus (56). The air change rate, $\lambda_a = 1.25$ ACH is estimated from previous studies of air quality in transit buses with closed windows (57). 3. The Diamond Princess cruise ship during its 12-day port quarantine in Yokohama, Japan (30, 31). We infer $I_0 = 20$ and $N\beta = 0.25$/day by fitting the confirmed case history to our fast-incubation solution, Eq. [S11], as shown in Fig. S1. We estimate relevant volume and area from floor plans of the Diamond Princess (33, 58), where passengers and crew mainly occupy 14 floors of living space of beam width 38 m, an average length equal to 90% of the ship's length 290 m, and mean ceiling height 2.1 m. We also assume a standard cruise-ship ventilation rate (8 ACH) for partially recirculated air conditioning (59). 4. Initial outbreak in Wuhan City, Hubei Province, China. We assume that the population-level spreading is dominated by indoor aerosol transmission with slow incubation (Eq. [S10]) in single-family apartments (60) with a mean family size of 3.03 (61), mean apartment area of 315 SF/person (62) and mean ventilation rate of 0.34 ACH (63). We estimate a mean exposure time of $\tau = 5.5$ days until symptoms (and patient isolation), and use the average $\mathcal{R}_0 = 3.3$ estimated for the population during the initial outbreak (64) in place of $\mathcal{R}_{in}(\tau)$.**

ting to the available data allows us to infer that the initial number of passengers infected on February 3, 2020 was approximately $I_0 = 20$, and that the concentration of infection quanta characterizing this particular spreading event was $C_q = 30$ quanta/$m^3$. Furthermore, it suggests that the incubation time is significantly less than 12 days, as is consistent with current estimates for the average time between exposure and the onset of symptoms being 2-5 days (54).

Our fitting of $I_0$ and $C_q$ for the Diamond Princess is based on the hypothesis of a "well-mixed ship". While such an approach is not traditional, and would be contested by those who do not believe that airborne transmission was prevalent on the Diamond Princess (68), it is consistent with a growing body of evidence (69). Analysis of the ship's floor plans and ventilation system has indicated recirculation of heated interior air throughout the entirety of the ship during cold weather (-5° C), without adequate filtration for virus-containing aerosol droplets (32, 33). Moreover, a detailed statistical analysis of the SARS-CoV-2 transmission history between passenger cabins revealed no significant correlation between new cases and the sharing of rooms with previously confirmed cases. Airborne transmission was further suggested by several examples of new cases emerging in single-occupancy cabins despite no known contact with other cases (32). Like many other indoor COVID-19 spreading events (53, 56, 70) in which position in an enclosed space was uncorrelated with likelihood of transmission (67), the cruise line outbreaks present evidence that strongly supports the notion of airborne transmission of SARS-CoV-2 through well-mixed indoor spaces (60, 71, 72).

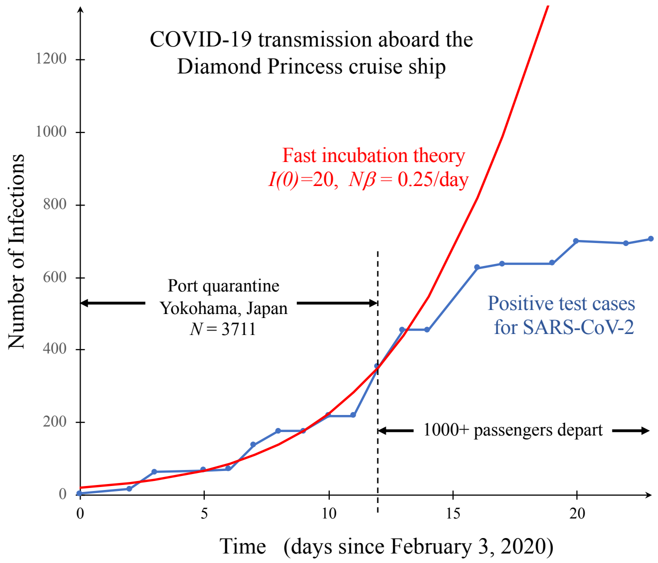


**Fig. S1.** Validation and calibration of the fast-incubation model for the super-spreading event on board the Diamond Princess cruise ship, during its twelve day quarantine at Yokohama, Japan in February 2020 (30). Fitting the number of SARS-CoV-2 positive cases versus time (blue data points) during the quarantine period, to the fast-incubation solution of the SEI model, Eq. [S11], (red curve) yields estimates for the initial number infected ($I_0 = 20$) and the transmission rate ($N\beta = 0.25$/day), from which we infer the concentration of infection quanta from breathing ($C_q = 30$ quanta/$m^3$). The relevant physical parameters are listed in Table S1.

**B. Ningbo tour bus.** We proceed by considering another super-spreading event, involving a Buddhist blessing ceremony held at the Tiantong Temple in Ningbo, Zhejiang Province, China on January 19, 2020 (55, 56). The confirmed index case, Ms. S, was thought to have contracted the virus two days earlier from dinner guests who had traveled to Wuhan during the initial outbreak. Ms. S traveled to the ceremony on a tour bus (Bus 2) for 50 minutes with 68 people, including the driver, and returned on the same bus with the same seating arrangement. Since it was winter, the windows were likely closed. No outside

| Activity | Experiment (74) | $Q_b$ (m$^3$/h) | $\phi_1$ ($10^{-16}$) | $C_q$ (q/m$^3$) | $\lambda_q$ (q/h) |
|---|---|---|---|---|---|
| breathing at rest | nose in, nose out (b-nn) | 0.5 | 0.35 | 8.8 | 4.2 |
| breathing heavily | nose in, mouth out (b-nm) | 0.5 | 1.3 | 33 | 16 |
| whispering | whispered counting (c-w-p) | 0.75 | 1.5 | 37 | 28 |
| speaking | voiced counting (c-v-p) | 0.75 | 2.9 | 72 | 54 |
| singing softly | whispered “aahs" (aah-w-p) | 1.0 | 4.1 | 103 | 103 |
| singing | voiced “aahs" (aah-v-p) | 1.0 | 39 | 970 (53) | 970 |

**Table S2. Activity dependence of airborne transmission of COVID-19. Expiratory droplet size distributions for different activities are taken from the experiments reported by Morawska et al. (74), and reasonable estimates are made for the exhaled air volume per time $Q_b$ from breathing (0.5 m$^3$/h), speaking (0.75 m$^3$/h) and singing (1.0 m$^3$/h). Our model is then used to predict the steady-state aerosol volume fraction, $\phi_1 = \int_0^{r_c} \phi_s(r)\, dr$, that results from exhalation of a single infectious individual in a setting corresponding to the Skagit Valley Choir room (Table S1), by integrating the distributions shown in Fig. 1 up to the critical radius $r_c = 2.5\mu$m. The concentration of COVID-19 infection quanta in the breath of an infected individual is assumed to be $C_q = 970$ q/m$^3$ for the singing case, as estimated for the Skagit Valley Chorale incident by Miller et al. (53), and values of $C_q$ for other activities are calculated by rescaling with the appropriate ratio of aerosol volume fractions, $\phi_1$. The quanta emission rate for each activity is then given by $\lambda_q = Q_b C_q$.**

air was supplied by mechanical ventilation, although the air was recirculated continuously. No masks were worn by the passengers, as there had not previously been any cases of COVID-19 in Zhejiang Province. Of the 68 passengers, 23 new cases of COVID-19 were confirmed, with their locations being evenly distributed across the bus, largely uncorrelated with proximity to Ms. S (56). Most had no close contact with the index case. However, some spatial patterns of transmission could be discerned on the bus, such as fewer cases among the window seats, possibly due to recirculation air flows. The epidemiological evidence for indoor airborne transmission is overwhelming (56).

Of the 172 others that attended the ceremony, only 7 individuals (4.1%) became infected at the 150-minute worship event, which was mostly held outside. Assuming that the bus passengers were infected at the temple at the same rate as the others, we may estimate that 3 of 23 transmissions occurred there, so only 20 on the round-trip bus ride. The published floorplan of the bus (56) resembles that of a Dongfeng 67-seat luxury tour bus made in Hubei Province, China, whose specifications we use to estimate interior dimensions. The most uncertain model parameter is the air exchange rate, $\lambda_a$, which we presume to be associated with natural ventilation. We further assume that windows were closed and that air filtration was limited. Although $\lambda_a = 1.8 - 3.7$/h has been measured in passenger cars with closed windows and recirculating fans (73), transit buses tend to have lower values, such as $\lambda_a = 1.25$/h (57), a value that we adopt here. We also assume a typical seated breathing flow rate, $Q_b = 0.5$ m$^3$/h.

Using these approximations for the model parameters, we apply the slow-incubation model (S10) to infer $C_q = 90$ q/m$^3$ and $\lambda_q = 45$ q/h. This value of $C_q$ falls in the range of intermediate speaking obtained from other inferences in the main text, which would be consistent with passengers speaking amongst themselves on the voyage. Given the uncertainty in the air exchange rate, the inferred value of $C_q$ could also be consistent with normal breathing with only occasional speech. Finally, we note that the average age of the bus passengers was 59 (56), which suggests a relatively vulnerable population, for which one expects to infer a relatively high $C_q$.

**C. Wuhan outbreak.** Table S1 also shows how the theory can be used to interpret the population-level reproductive number in terms of indoor airborne transmission, assuming that the primary disease transmission arises in family apartments, as shown in a recent analysis of COVID-19 spreading in China (60). We consider the initial outbreak in Wuhan, Hubei Province and assume physical parameters appropriate for typical apartments and families. We then use the average population reproductive number (64) to assert $\mathcal{R}_{in}(\tau) = \langle \mathcal{R}_0 \rangle = 3.3$, where $\tau = 5.5$ days is the mean time for an newly infected family member to show symptoms and be isolated or removed.

As shown in the main text (Fig.2), the inferred values of $C_q = 29$ quanta/m$^3$ for family apartments in the Wuhan outbreak and $C_q = 30$ quanta/m$^3$ for the Diamond Princess cruise ship are consistent with the light respiratory activities expected to be most prevalent in those settings, such as sleeping and quiet speech. Both inferred $C_q$ values are greatly exceeded by the inference of $C_q = 870$ (made here, using $\bar{r} = 2.0\mu$m and $\lambda_a = 0.65$/h; see Table S1) or 970 quanta/m$^3$ (made by Miller et al (53), averaging over simulations with variable $\lambda_a = 0.3-1.0$/h and $\lambda_s = 0.3-1.5$/h) for singing, but in a manner consistent with the increased pathogen output associated with more vigorous speaking or singing (23, 75). These inferences build confidence in our estimates of exhaled quanta concentrations, $C_q$, for various respiratory activities, as shown in Figure 2 of the main text.

### 6. The dependence of airborne disease transmission on respiratory activity

It is well established that aerosol droplet production varies strongly with the form of expiratory activity (74, 76). For example, vocalization greatly increases aerosol emission relative to quiet breathing, roughly in proportion to the amplitude of the sound produced (23, 77). In Table S2, the aerosol volume fraction produced by different activities, obtained by integrating the distributions in Fig. 1 up to $r_c = 2.5\mu$m, are used to rescale the inferred quanta concentration for singing (53) in the Skagit Valley Choir room into those appropriate for other activities. The resulting values are consistent with the values of $C_q$ inferred for the Diamond Princess quarantine and the initial Wuhan City outbreak (see Table S1 and Fig. 2).

### 7. Application of the Safety Guideline

In order to illustrate how to implement our safety guideline, we provide a spreadsheet as Supplementary Data, which has also been implemented for general use as an online app, translated into many languages (78). We note that, following the submission of our manuscript, it has come to our attention that the groups of José-Luiz Jimenez (79) and Lidia

Morawska (80) have produced spreadsheets with extensive documentation (17, 81) similar in spirit to ours, designed to provide quantitative risk assessments for safe management of indoor spaces during the COVID-19 pandemic. Since their analyses and others (82, 83) are based on the same physical picture of airborne transmission in a well-mixed space, their predictions are consistent with ours. However, their recommendations rely on calculations of pathogen concentration and risk in different settings, while ours are based entirely on our Safety Guideline, as succinctly expressed by the bound on cumulative exposure time in Eq. [5].

Our spreadsheet and online app enable the calculation of the suggested maximum cumulative exposure times for specific indoor spaces. In this section, we offer guidance on how to use the spreadsheet for both safety assessment and contact tracing, specifically, how to select suitable parameters and properly interpret the results. The input parameters, colored in pink in the spreadsheet and in *italics* below, are divided into the following four categories.

**A. Physical Parameters..** The geometry of the indoor space is specified by its *floor area*, $A$, and *mean ceiling height*, $H$, from which the volume $V = AH$ is calculated using appropriate unit conversions. The *ventilation outflow rate*, $Q = V\lambda_a$, is calculated from the air exchange rate, $\lambda_a$, typically expressed in terms of air changes per hour (ACH). This critical input parameter is governed by national or local standards for different types of indoor spaces, such as the ASHRAE standards (62.1) in the United States (84). As noted in the main text, natural ventilation may be approximated as $\lambda_a = 0.34$/h, which has been measured in bedrooms with closed windows (63) and is considered to be the minimum standard (84), although this value will vary with both location and quality of construction. For residences, classrooms, businesses, and public spaces, $\lambda_a$ usually falls in the range $4-8$/h. Crowded spaces, such as bars, nightclubs and restaurants, typically require more vigorous ventilation, $\lambda_a = 15 - 20$/h. Minimum ventilation standards for American hospitals have increased from 12 to 18 ACH (85). Most chemical and biological laboratories have $\lambda_a$ in the range of $6 - 12$/h, but those handling toxic or infectious materials may have $\lambda_a$ as high as $20 - 30$/h. Revised ASHRAE standards, intended to mitigate the spread of airborne infectious diseases, recommend a minimum of $\lambda_a = 6$/h for all indoor spaces (86).

Forced air filtration and droplet settling in ducts may also be included through the effective air filtration rate, $\lambda_f(r) = p_f(r)\lambda_r$, where $\lambda_r = Q_r/V$ is the *recirculation air exchange rate* passing through the filter and $p_f(r)$ is *droplet filtration probability*, taken to be a constant over the aerosol size range. The United States Environmental Protection Agency defines high-efficiency particulate air (HEPA) filtration (87) as removing $p_f = 99.97\%$ of aerosol particles, while ordinary air filters are assigned Minimum Efficiency Reporting Value (MERV) ratings corresponding to $p_f > 20\%$–$90\%$ in specified aerosol size ranges (88). For ventilation systems with indoor air recirculation, the primary outdoor air fraction, $Z_p = Q/(Q + Q_r)$, is usually specified by indoor air quality (IAQ) standards. For example, $Z_d = 20\%$ for classrooms in the United States (89)), where $Q + Q_r$ is the total flow rate and $v = (Q+Q_r)/A$ is the mean air velocity. Alternatively, air filtration may be accomplished by indoor free-standing units with a specified recirculation flow rate of $Q_r$.

The *relative humidity* $RH$ of the indoor air is another physical input parameter, which affects both respiratory droplet evaporation and viral deactivation (90, 91).

**B. Physiological Parameters.** The first physiological parameter is the *volumetric breathing flow rate*, $Q_b$, which is approximately 0.5 $m^3$/h for resting and light activity. Average values for healthy males and females have been reported as 0.49, 0.54, 1.38, 2.35, and 3.30 $m^3$/h for resting, standing, light exercise, moderate exercise and heavy exercise, respectively (92), and used in simulations of airborne transmission of COVID-19 (16). The second physiological parameter is the mean respiratory aerosol droplet size $\bar{r}$ for the suspended infectious droplets responsible for airborne disease transmission. The precise definition of $\bar{r}$ is given in Eq. (S7), in terms of the distribution of droplet sizes for different types of respiration (23, 74, 77), the size-dependent infectivity of aerosol droplets (65), and the settling and ventilation rates. As illustrated in the main text, a typical value for $\bar{r}$ is 2-3 $\mu$m. We note that these values are roughly consistent with the standard definition of aerosol droplets, as those having $r < 5\mu$m (16). The effect of relative humidity on the size of stable droplets after evaporation (93) can be estimated by rescaling $\bar{r}$ by $\sqrt[3]{0.4/(1 - RH)}$, since the droplet distributions used to calibrate the guideline were measured at $RH = 60\%$ (74).

**C. Disease Parameters.** The most important disease parameter is the *infectiousness of exhaled air*, $C_q$, the infection quanta per volume. Using all of the limited information currently available, we estimate $C_q = 30$ q/$m^3$ for normal breathing and light activity and provide our best estimates of $C_q$ for different respiratory activities in Fig. 2. Our analysis indicates that $C_q$ can be an order of magnitude larger for singing or other vigorous respiratory activities, or an order of magnitude smaller for sleeping and light nose breathing.

The second disease parameter, $s_r$, is the *relative transmissibility* (or susceptibility) of the virus. This transmissibility necessarily depends not only on the susceptibility of the population, which is known to be age-dependent (54, 94–98), but on the viral strain (99, 100). For example, a study of transmission in quarantined households during lockdowns in China reported $s_r = 0.23$ for children (aged 0–14) and $s_r = 0.68$ for adults (aged 15-64) relative to $s_r = 1$ for the elderly (over 65 years old) (101). Initial estimates of $\mathcal{R}_0$ from the United Kingdom for the new variant of concern (VOC 202012/01, lineage B.1.1.7) suggest that $s_r$ is increased by 60% relative to the original strain of SARS-CoV-2 and show signs of elevated risk of infection among children (99, 100).

The third disease parameter is the *viral deactivation rate*, $\lambda_v$, at which the aerosol-bound virus loses infectiousness, which for SARS-CoV-2 has been estimated to lie in the range of zero (66) to 0.63/h (67). Taking into account results for other aerosolized viruses (90, 91, 102), the viral deactivation rate is approximated as linear in relative humidity, $\lambda_v = \lambda_{v,50}RH$, where the value $\lambda_{v,50}$ is specified. The effective viral deactivation rate may also be enhanced by ultraviolet radiation (UV-C) (103) or airborne dispersal of chemical disinfectants (*e.g.* $H_2O_2$, $O_3$) (104).

**D. Precautionary Parameters.** The first precautionary parameter is the *mask penetration factor*, $p_m$, defined as the fraction of infectious aerosol droplets that pass through the mask during exhalation or inhalation, averaging over any dependencies on respiratory activity (105) or direction of airflow (106). Alternatively, the *filtration efficiency* is defined as $1 - p_m$. Many studies are available to help assign this value for different types of face coverings, ranging from cloth coverings to surgical masks (107–110). Although filtration efficiency depends on drop size, it is typically nearly constant in the aerosol range, up to several microns in diameter (107, 108). Typical values for disposable medical masks are in the range $p_m = 1-5\%$ and vary with fit and applied pressure (108, 109), while for simple cloth face masks, values of $p_m = 10 - 40\%$ are typical for hybrid, multi-layer fabrics and $p_m = 40 - 80\%$ for single-layer or coarse fabrics (110).

The second precautionary parameter is the *disease transmission tolerance*, $\epsilon$. We note that $\epsilon = 1$ corresponds to the baseline of one expected transmission during the occupancy period. The choice of $\epsilon$ should take into account the vulnerability of the population, which for COVID-19 is a strong function of age and pre-existing medical conditions (54, 95, 111). Relative to the median age of 69 in the Skagit Valley Chorale spreading incident used to calibrate our model, the relative rate of hospitalization with COVID-19 (95) can be calculated as 2.5% (ages 0-4), 0.8% (ages 5-17), 20% (ages 18-49), 61% (ages 50-64), 130% (ages 75-84), and 145% (ages $> 85$). For the elderly, especially those with preexisting conditions or co-morbidity, $\epsilon \ll 1$ should be chosen. For the young and healthy (in regions where hospitals are not overwhelmed and vulnerable groups are protected), larger values of $\epsilon$ could be considered (112). As noted in the main text, choosing a sufficiently small $\epsilon$ will also serve to mitigate against prolonged exposure to respiratory jets, whose contribution to pathogen transport may dominate in the absence of face-mask use (3).

**E. Results.** The spreadsheet first computes properties of the infectious aerosol per infected individual in the room, which are primarily of technical interest: the effective droplet settling speed $v_s(\bar{r})$, the concentration relaxation rate $\lambda_c(\bar{r})$, the dilution factor, $f_d$, and the infectiousness of the ambient air, $f_d C_q$, in steady state per infected person in the room.

The spreadsheet computes the safety guideline in two ways with the key results highlighted in green. First, the occupancy limit $N_{max}$ can be calculated for a given exposure time $\tau$, as is set by the typical residence time of people in the indoor space. The corresponding minimum outdoor airflow per person, $Q/N_{max}$, may be compared with local standards, such as 3.8 L/s/person for retail spaces and classrooms and 10 L/s/person for gyms and sports facilities in Europe (113), or the ASHRAE Standards in the United States, typically 5-20 cfm/person depending on the type of space (84). Second, the time limit $\tau_{max}$ is calculated for a given occupancy $N$. These bounds are plotted and may be compared to the Six-Foot Rule and 15-Minute Rule, both of which invariably violate our guideline. For the bounds on both $N_{max}$ and $\tau_{max}$, two results are reported: the transient bound, which accounts for the buildup of infectious aerosols in the air after the entrance of an infected person, and the more conservative steady-state bound, which is relevant after the relaxation time $\lambda_c \tau \gg 1$.

**F. Contact Tracing.** The spreadsheet can also be used as a tool for contract tracing. With a conservative tolerance, such as $\epsilon = 0.01$, the guideline defines whether or not the $N$ occupants of a room visited by the index case for a time $\tau$ should be considered as contacts for the purpose of tracing the infection network. If the guideline is violated, then *all* occupants of the room must be considered contacts, regardless of their distance from the index case. Compared to the current CDC definition of contact (114) – spending more than 15 minutes less than 6 feet apart from an infected person – this definition, based on our consideration of airborne transmission, may thus identify significantly more contacts to be traced and quarantined.

**G. Disclaimer.** Our Indoor Safety Guideline calculator is an evolving tool intended to familiarize the interested user with the factors influencing the risk of indoor airborne transmission of COVID-19, and to assist in the quantitative assessment of risk in various settings. We note that uncertainty in and intrinsic variability of model parameters may lead to errors as large as an order of magnitude, which may be compensated for by choosing a sufficiently small risk tolerance. Our guideline does not take into account short-range transmission through respiratory jets, which may substantially elevate risk when face masks are not being worn, in a manner discussed in the main text. Use of the Indoor Safety Guideline is the sole responsibility of the user. It is being made available without guarantee or warranty of any kind. The authors do not accept any liability from its use.

### 8. Online course

The scientific principles underlying this work are also taught in a free, self-paced massive, open online course (MOOC) (115).

### 9. Glossary of Mathematical Symbols

See Table S3.

| Symbol | Meaning | Characteristic values |
|---|---|---|
| Engineering Parameters | | |
| $N$ | Number of persons, room occupancy | 1 – 1000 |
| $\tau$ | Time since an infected person entered the room | 0-1000 h |
| $V$ | Room volume | 10–$10^4$ $m^3$ |
| $A$ | Floor surface area | 5–5,000 $m^2$ |
| $H$ | Mean ceiling height, $V/A$ | 2–6 m |
| $Q$ | Ventilation outflow rate | 1–$10^5$ $m^3$/h |
| $\lambda_a$ | Ventilation (outdoor air exchange) rate, $Q/V$ | 0.1–30 $h^{-1}$ |
| $Q_r$ | Recirculation flow rate | 1–$10^5$ $m^3$/h |
| $\lambda_r$ | Recirculation air exchange rate, $Q_r/V$ | 0.1–30 $h^{-1}$ |
| $Z_p$ | Primary outdoor air fraction, $Q/(Q+Q_r)$ | 0.1-1.0 |
| $p_f$ | Probability of droplet filtration via recirculation | 0–1.0 |
| $\lambda_f(r)$ | Filtration removal rate, $p_f\lambda_r$ | 0–30 $h^{-1}$ |
| Physical Parameters | | |
| $r$ | Respiratory drop radius | 0.1–100 $\mu$m |
| $r_c$ | Critical airborne drop radius | 0.2–20$\mu$m |
| $V_d$ | Drop volume, $\approx \frac{4}{3}\pi r^3$ | $10^{-5}$–$10^6$$\mu m^3$ |
| $n_d(r)$ | Drop number density per radius | 0.01–1.0 drops/($cm^3 \cdot \mu$m) |
| $\rho_a$ | Air density | 1.2 kg/$m^3$ |
| $\rho_d$ | Drop mass density | 1000–1500 kg/$m^3$ |
| $\Delta\rho_d$ | Drop relative density, $\Delta\rho = \rho_d - \rho_a$ | 1000–1500 kg/$m^3$ |
| $\mu_a$ | Air viscosity | 18 $\mu$Pa·s |
| $g$ | Gravitational acceleration | 9.8 m/$s^2$ |
| $v_s(r)$ | Drop settling speed | $10^{-5}$–$10^2$ mm/s |
| $\lambda_s(r)$ | Drop settling rate, $v_s(r)/H$ | $10^{-5}$–$10^2$ $h^{-1}$ |
| $\phi_s$ | Drop initial solute volume fraction | 0.01–10% |
| $RH$ | Relative humidity | 10–100% |
| $r_{eq}$ | Equilibrium drop radius after evaporation | $10^{-2}$-10$\mu$m |
| $\alpha_t$ | Turbulent entrainment coefficient of respiratory jet | 0.1–0.15 |
| $M$ | Momentum flux of a respiratory jet | $10^3$ - $10^4$ $cm^4/s^2$ |
| $A_m$ | Average area of the mouth opening | 1-10 $cm^2$ |
| $C_j(x)/C_0$ | Turbulent pathogen concentration in a jet relative to the mouth | 0–1 |
| $x$ | Distance from infected person | 0.1-10 m |
| $p_j$ | Probability of being in the respiratory jet of an infected person | 0–1 |
| Epidemiological Parameters | | |
| $S(t)$, $E(t)$, $I(t)$, $R(t)$ | Number of susceptible, exposed, infected and recovered persons | |
| $\alpha$, $\beta$, $\gamma$ | Incubation, transmission and removal rates | |
| $\beta_a(t)$ | Mean airborne transmission rate per infected/susceptible pair | $10^{-6}$–10 quanta/$h^{-1}$ |
| $\bar{\beta}_a$, $\langle\beta_a\rangle$ | Steady-state and time-averaged mean transmission rate | $10^{-6}$–10 quanta/$h^{-1}$ |
| $Q_b$ | Mean breathing flow rate | 0.5–3.0 $m^3$/h |
| $\lambda_v$ | Pathogen (virion) deactivation rate | 0.01–10 $h^{-1}$ |
| $\lambda_c(r)$ | Pathogen concentration relaxation rate | 0.1-100 $h^{-1}$ |
| $\bar{r}$ | Effective infectious drop radius | 0.3–5$\mu$m |
| $f_d$ | Pathogen dilution factor, $Q_b/(\lambda_c(\bar{r})V)$ | $10^{-5}$–$10^{-2}$ |
| $P(r)$ | Pathogen production rate per air volume per drop radius | $10^{-6}$–$10^9$ virions/(h·$\mu$m) |
| $C(r,t)$ | Mean pathogen concentration per air volume per drop radius | $10^{-8}$–$10^4$ virions/($m^3 \cdot \mu$m) |
| $C_s(r)$ | Steady-state airborne pathogen concentration per drop radius, $P(r)/(\lambda_c(r)V)$ | $10^{-8}$–$10^4$ virions/($m^3 \cdot \mu$m) |
| $\phi_s(r)$ | Steady-state infectious aerosol volume fraction distribution, $C_s(r)/c_v(r)$ | $10^{-19}$–$10^{-13}/\mu$m |
| $\phi_1$ | Steady-state aerosol volume fraction (from one infected person), $\int_0^{r_c}\phi_s(r)dr$ | $10^{-19}$–$10^{-12}$ |
| $c_v(r)$ | Pathogen (virion) concentration per drop volume | $10^4$–$10^{11}$ RNA copies/mL |
| $c_i(r)$ | Pathogen infectivity, $1/$(infectious dose) | 0.001–1.0 |
| $c_q(r)$ | Concentration of infection quanta per drop volume, $c_i c_v$ | 10–$10^{11}$ quanta/mL |
| $C_q$ | Infectiousness of breath, exhaled quanta concentration | 1–1000 quanta/$m^3$ |
| $\lambda_q$ | Quanta emission rate, $Q_b C_q$ | 1–1000 quanta/h |
| $p_m(r)$ | Probability of drop transmission through mask, $1-$(filtration efficiency) | 0.01–0.1 |
| $p_i$ | Probability a person is infected, prevalence of infection | 0–1 |
| $p_s$ | Probability a person is susceptible (not exposed, immune, or vaccinated) | 0–1 |
| $s_r$ | Relative transmissibility (or susceptibility) | 0.1–10 |
| $N_s$ | Expected number of susceptible persons $\leq N$ | 0–1000 |
| $\mathcal{R}_0$ | Reproductive number, $S(0)\beta/\gamma$ | 0.01–10 |
| $\mathcal{R}_{in}$ | Indoor reproductive number, $N_s s_r \langle\beta_a\rangle\tau$ | 0.001–100 |
| $\epsilon$ | Risk tolerance, bound on expected number of transmissions | 0.005–0.5 |
| $N_{max}$ | Maximum safe room occupancy | 1–1000 |

**Table S3. Terms arising in our theoretical formulation, their units and characteristics values.**
